# Cilostazol and isosorbide mononitrate for the prevention of progression of cerebral small vessel disease: Baseline data and statistical analysis plan for the Lacunar Intervention Trial-2 (LACI-2) (ISRCTN14911850)

**DOI:** 10.1101/2022.05.31.22275743

**Authors:** Philip M Bath, Iris Mhlanga, Lisa J Woodhouse, Fergus Doubal, Katherine Oatey, Alan A Montgomery, Joanna M Wardlaw, the LACI-2 Investigators

**Affiliations:** Stroke Trials Unit, Mental Health & Clinical Neuroscience, University of Nottingham, Nottingham UK; Stroke, Nottingham University Hospitals NHS Trust, Nottingham UK; Neuroimaging Sciences, Centre for Clinical Brain Sciences, University of Edinburgh, Edinburgh UK; Edinburgh Clinical Trials Unit, University of Edinburgh, Edinburgh UK; Nottingham Clinical Trials Unit, University of Nottingham, Nottingham UK; UK Dementia Research Institute at the University of Edinburgh

**Keywords:** cerebral small vessel disease, cilostazol, isosorbide mononitrate, lacunar stroke, statistical analysis plan, randomised clinical trial

## Abstract

**Background:** Cerebral small vessel disease (SVD) causes lacunar strokes (25% of all ischaemic strokes), physical frailty and cognitive impairment and vascular and mixed dementia. There is no specific treatment to prevent progression of SVD.

**Methods:** The LACunar Intervention Trial-2 (LACI-2) is an investigator-initiated prospective randomised open-label blinded-endpoint (PROBE) phase II feasibility study assessing cilostazol and isosorbide mononitrate for preventing SVD progression. We aimed to recruit 400 patients with clinically-evident lacunar ischaemic stroke and randomised to cilostazol, isosorbide mononitrate, both or neither, in addition to guideline secondary ischaemic stroke prevention, in a partial factorial design. The primary outcome is feasibility of recruitment and adherence to medication; key secondary outcomes include: drug tolerability; recurrent vascular events, cognition and function at one year after randomisation; and safety (bleeding, falls, death). Data are number (%), and median [interquartile range].

**Results:** The trial commenced on 5^th^ of February 2018 and ceased recruitment on 31^st^ of May 2021 with 363 patients randomised, with the following baseline characteristics: average age 64 [56.0, 72.0] years, female 112 (30.9%), stroke onset to randomisation 79.0 [27.0, 244.0] days, hypertension 267 (73.6%), median blood pressures 143.0 [130.0, 157.0]/83.0 [75.0, 90.0] mmHg, current smokers 67 (18.5%), educationally achieved end of school examinations (A-level) or higher 118 (32.5%), modified Rankin scale 1.0 [0.0, 1.0], National Institutes Health stroke scale (NIHSS) 1.0 (1.4), Montreal Cognitive Assessment 26.0 [23.0, 28.0] and total SVD score on brain imaging 1.0 [0.0, 2.0]. This publication summarises the baseline data and presents the statistical analysis plan.

**Summary:** The trial is currently in follow-up which will complete on 31 May 2022 with results expected in October 2022.

## INTRODUCTION

Cerebral small vessel disease (SVD) is a major cause of stroke (lacunar ischaemic stroke), intracerebral haemorrhage and vascular and mixed dementias.^1^ It is most commonly due to an intrinsic disorder of the brain’s small perforating arterioles with endothelial dysfunction manifesting as impaired vasoreactivity, vascular stiffness, blood–brain barrier leakage and perivascular inflammation.^1^ Currently, there is no specific secondary prevention for lacunar ischaemic stroke or SVD-associated cognitive decline.^2^ Combined aspirin and clopidogrel increased bleeding ^3^ and intensive antihypertensive therapy did not reduce recurrence or prevent cognitive decline ^4^ in lacunar ischaemic stroke.

However, there are other potential interventions that might prevent SVD or reduce its development and progression, including: drugs that modulate the blood brain barrier or lower blood lipids, immunosuppressive agents, neurotrophins, peroxisome proliferator-activated receptor-gamma agonists, rho-kinase antagonists, vitamins, anti-inflammatory agents and xanthine oxidase inhibitors.^2 5^ Additionally, drugs that modulate the nitric oxide (NO)-cyclic guanylate cylase-phosphodiesterase-5 and prostacyclin (PGI_2_)-cyclic adenylate cylase-phosphodiesterase-3 systems are attractive since they mimic key endogenous vaso-regulators.^2^ Both NO and PGI_2_ have vasodilator, antiplatelet, antileucocyte,^6^ anti-smooth muscle and pro-endothelial-effects. NO may be administered orally as substrate (L-arginine or inorganic nitrate), organic nitrates (such as isosorbide mononitrate, ISMN) or drugs that inhibit PDE5 (such a dipyridamole ^7^ or sildenafil) to increase and preserve levels of the NO second messenger, cyclic guanylate monophosphate. Although PGI_2_ has to be administered intravenously, PDE3 may be inhibited with cilostazol ^8^ which preserves levels of the PGI_2_ second messenger, cyclic adenylate monophosphate; cilostazol has showed promise in secondary prevention trials in ischaemic stroke, which included large proportions of patients with lacunar stroke subtype;^8^ and is widely used in east-Asia. In addition, in experimental models, cilostazol reversed impaired oligodendrocyte precursor cell maturation which occurs when endothelial cells are dysfunctional thus potentially reducing myelin damage and facilitating its repair,^9^ and increased the astrocyte-to-neuron lactate shuttle thus increasing energy supply and prolonging neuronal survival.^10^ Both actions are potentially relevant to reducing brain damage in SVD.

SVD can present clinically as lacunar ischaemic stroke, cognitive impairment, mobility and/or mood disorders, or be covert and detected on incidentally on brain imaging.^1^ These presentations are associated with increased short and long-term risk of recurrent stroke, cognitive decline, dementia, and functional impairments.^11-15^ However, although these clinical outcomes are highly relevant to patients and clinical services,^15 16^ the data on their rates long-term, individually or in combination, are sparse, precluding reliable sample size estimations for clinical trials in SVD.^17^ Furthermore, it was unclear if patients with clinically-evident lacunar ischaemic stroke could be identified with key prognostic variables determined accurately in routine clinical practice, or recruited in sufficient numbers rapidly enough, to make a clinical trial in stroke presentations of SVD feasible in a practical time-period. This is because lacunar ischaemic stroke may be confused clinically with mild cortical ischaemic stroke,^18^ MRI is not always available acutely nor positive for small subcortical stroke,^19^ and long-term outcomes vary with severity of SVD lesions on brain imaging,^13-15^ making it necessary to minimise the randomisation on SVD severity.^20^ Since lacunar ischaemic stroke is currently managed according to guidelines covering the whole of secondary prevention of ischaemic stroke (representing best medical care) and there is no justification currently for withholding secondary prevention, any novel drugs should be tested against a background of guideline secondary ischaemic stroke prevention.

The LACI-1/2 series of trials are assessing the feasibility of recruiting patients with clinically-evident lacunar ischaemic stroke and tolerability of treatment with ISMN and cilostazol,^17 21-24^ while also gathering data on outcome event rates, with the aim of testing these agents in a large phase III efficacy trial. In LACI-1, short-term administration of cilostazol and ISMN in addition to best medical care were well tolerated when the dose was escalated, without safety concerns, in patients with clinically-evident lacunar ischaemic stroke;^23^ additionally, these drugs appeared to reduce arterial stiffness without effects on platelet function ^22^ and appeared to improve cerebrovascular function.^24^

The present trial, LACI-2, is testing the long-term feasibility and tolerability of administering cilostazol and ISMN to patients with clinically-evident lacunar ischaemic stroke given on top of guideline secondary ischaemic stroke prevention while assessing safety and gathering data on clinically-relevant outcome event rates.^17^ Here we present the statistical analysis plan (SAP) and baseline data of LACI-2.

## METHODS

The LACunar Intervention Trial-2 (LACI-2) is a UK-based investigator-initiated, prospective, randomised, partial factorial, open-label, blinded-endpoint (PROBE) phase II feasibility trial of cilostazol and/or isosorbide mononitrate. Patients with clinically-evident lacunar ischaemic stroke, with no limit on the time interval since the stroke, with capacity to consent and who were independent in activities of daily living, were randomised to cilostazol, isosorbide mononitrate, both or neither in a partial factorial design. Patients with contraindications to one of the trial drugs could be randomised to the other drug alone. Randomised treatment was given in all patients in addition to guideline stroke prevention therapies, typically clopidogrel or aspirin, antihypertensive drugs and statins. Patients requiring oral anticoagulation were excluded.

Full details of regulatory approvals, patient assessment, inclusion and exclusion criteria, randomisation, minimisation criteria (age, sex, stroke severity [NIHSS], dependency resulting from the stroke [mRS], systolic blood pressure, smoking status, time after stroke, and highest level of education), dose escalation scheme, short and long term assessments are given in the protocol paper.^17^

The presence of an infarct relevant to the clinical presentation and a simplified estimate of total SVD lesion severity score, adapted for use on CT or MRI and by people not expert in brain imaging of SVD,^17 21^ were determined at the recruiting site. All diagnostic CT or MRI, scans obtained at any stroke recurrence, and one year follow-up MRI, were collected by the trial office for central adjudication.

The primary outcome is feasibility of recruitment and adherence to medication. Key secondary outcomes include recruitment of hospital sites and participants, drug tolerability, symptoms (such as headache, nausea, palpitations) which might deter drug adherence, safety (bleeding, falls, death), recurrent vascular events (including stroke and myocardial infarction), cognition and function over one year of follow-up. Imaging outcomes (white matter hyperintensity [WMH] severity, new cortical or subcortical infarcts, lacunes, haemorrhage, microbleeds) are assessed with magnetic resonance imaging (MRI) at one year. Details of central CT and MRI adjudication are given in the LACI-2 protocol paper.^17^

LACI-2 will provide outcome event rates and hence information to estimate sample size, recruitment and trial procedures for a phase III trial based in clinical outcomes. Further information is given in the Supplement and published protocol.^17^

We present here a complete listing of baseline data; data are number (%), and median [interquartile range], unless otherwise stated.

The full details of the Statistical Analysis Plan (SAP) ^25 26^ are given in the accompanying Supporting Information Appendix S1 and is presented prior to locking of the study database so that analyses are not data driven or reported selectively.^27^ The SAP also lists planned secondary analyses and substudies, and follows the layout suggested in guidelines.^25 26^

## RESULTS

The trial commenced in February 2018; recruitment was suspended due to COVID-19 restrictions on 17 March 2020 by the Sponsor and reopened on 10 June 2020; however the additional delays in reinstating site approvals meant that there was a total of four months without recruitment. Recruitment will stop on 31 May 2021 due to financial constraints, a total of 40 months (36 months excluding COVID-19 suspension).

Of a planned 400 participants, 363 (91%) were recruited from 26 UK hospitals. At baseline, the average age was 64 (range 31-87) years. There were 112 (31%) female patients, the median NIHSS was 0 [0, 0], the modified Rankin scale (mRS) was 2 in 85 (23%) patients, and the time from stroke onset to randomisation was 79.0 [27.0, 244.0] days (Table 1). The median age for completing education was 16 years, with 118 (33%) participants having one or more A-level equivalent or above qualifications. Vascular risk factor rates included median blood pressure of 143.0 [130.0, 157.0]/83.0 [75.0, 90.0] mmHg, current smoking in 67 (19%), drug-treated hypertension in 258 (71%), drug-treated hyperlipidaemia in 278 (77%), diabetes mellitus in 80 (22%), atrial fibrillation in 5 (1%), carotid stenosis of >50% left 3 (0.8%), right 6 (1.7%) and history of previous stroke in 25 (7%).

**Table 1.**
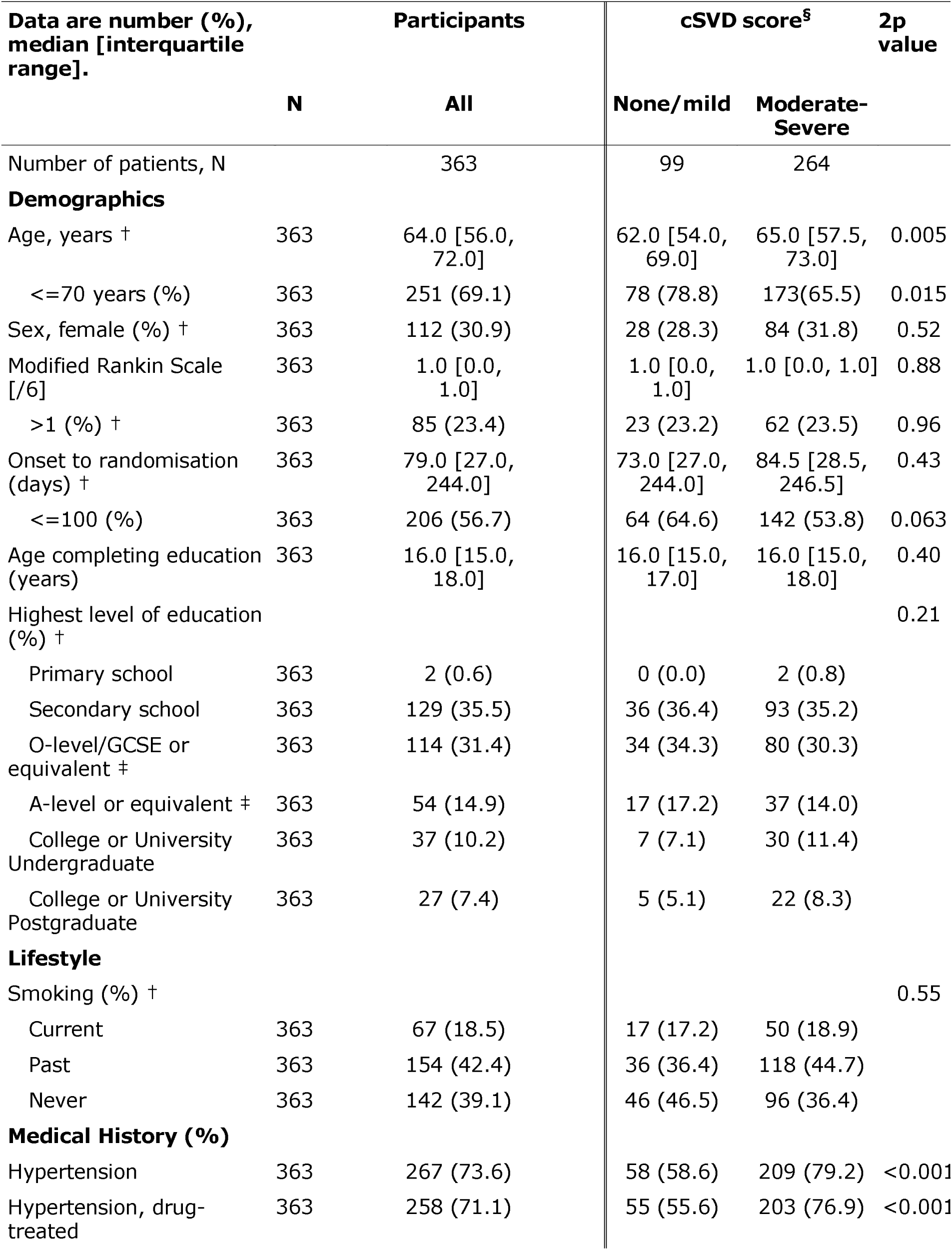
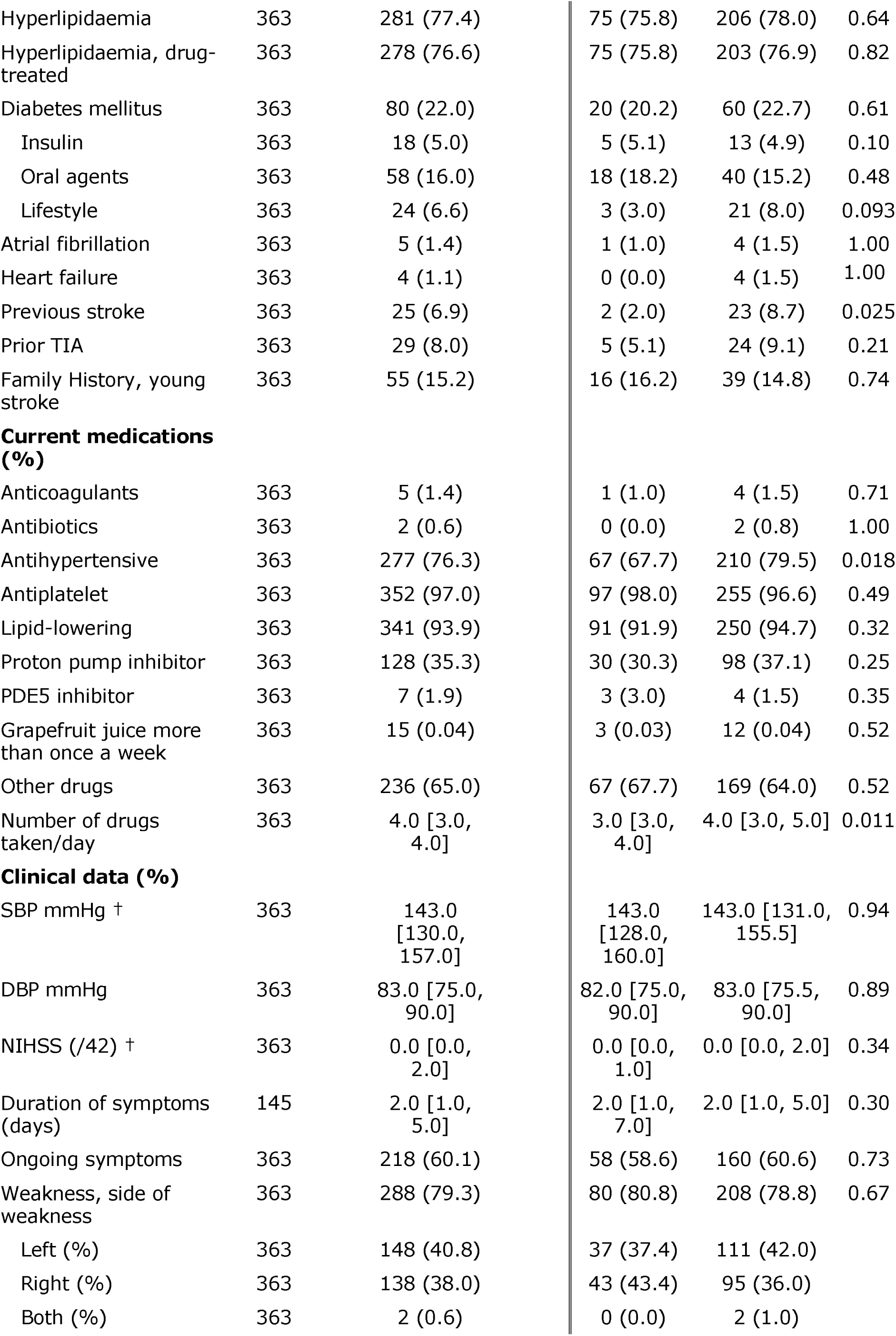
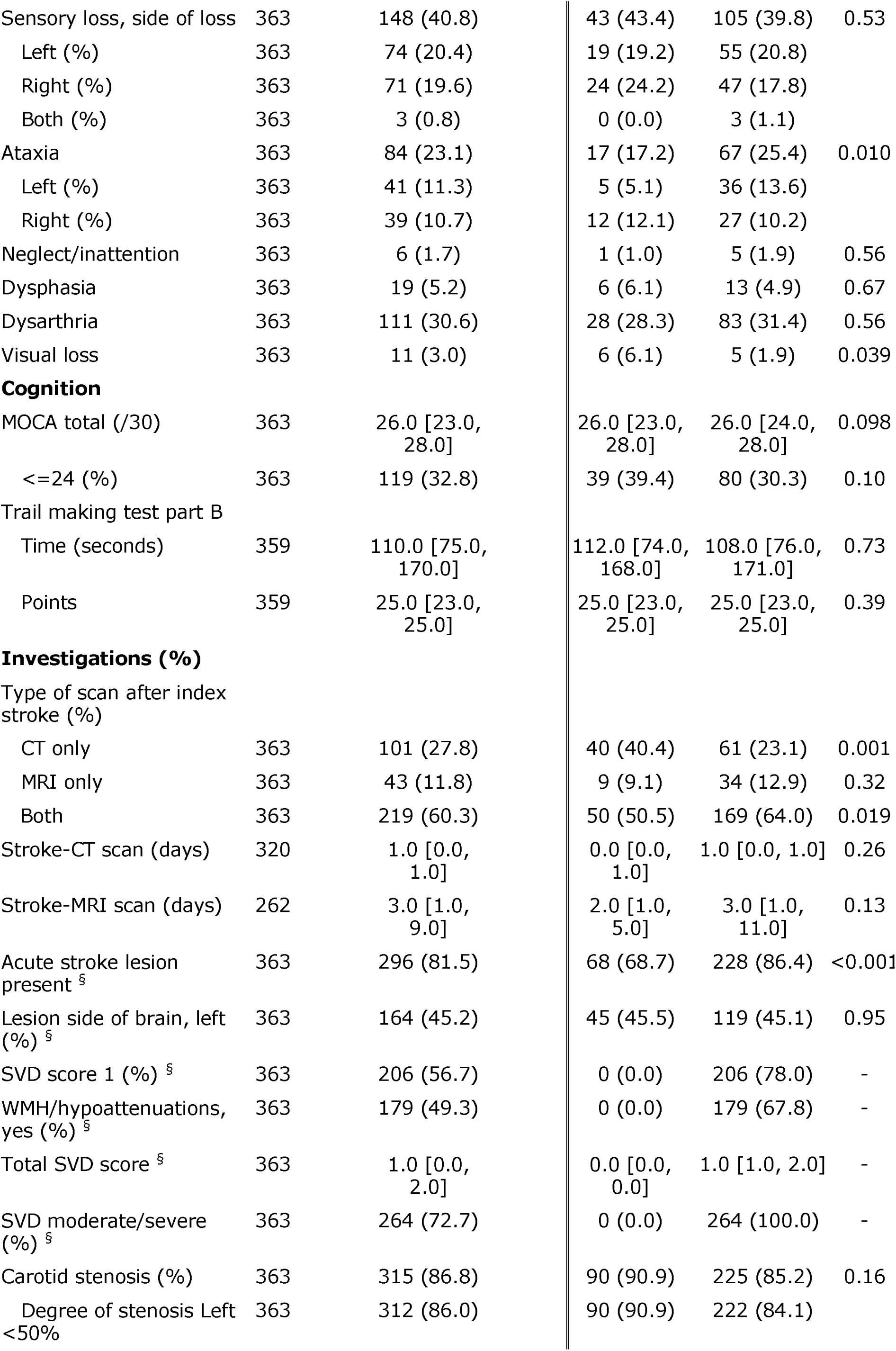
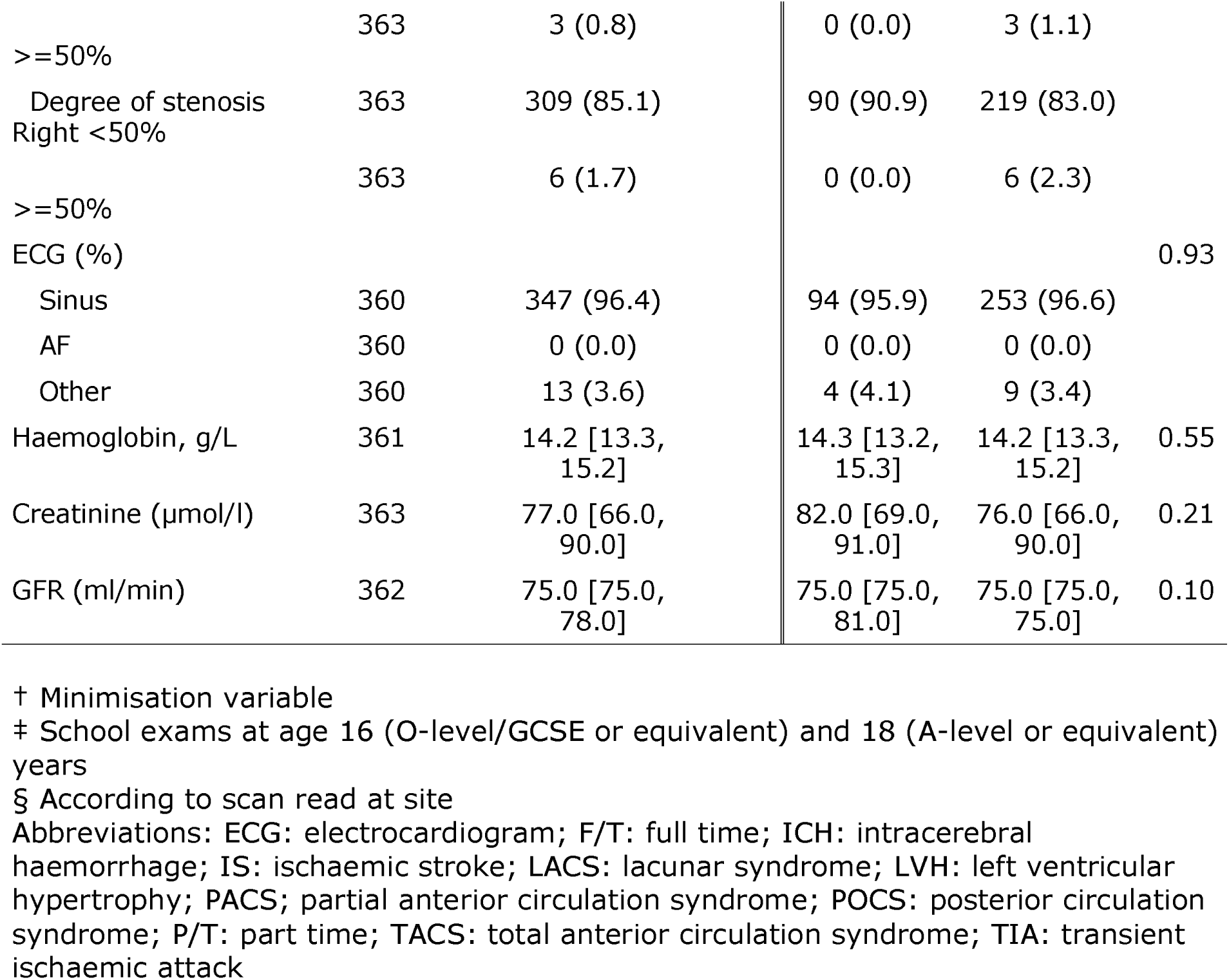
Baseline characteristics by small vessel disease score as determined by recruiting site.

Baseline clinical findings included persistent unilateral weakness in 288 (79%), sensory change in 148 (41%), neglect/inattention in 6 (2%), dysphasia in 19 (5%) and visual disturbance in 11 (3%) (Table 1). On cognitive testing, the median Montreal Cognitive Assessment score was 26.0 [23.0, 28.0] with 119 (33%) having a score below 25 signifying cognitive impairment. The median score on the trail making test, part B, a test of executive function, was 25.0 [23.0, 25.0] points.

A majority of participants, 219 (60%), had both an admission CT scan and MRI brain imaging (320, 88%, had CT and 229, 65%, had MRI). Imaging findings as reported by the recruiting site included an acute ischemic stroke lesion thought to be consistent with the lacunar stroke symptoms in 296 (82%), white matter hyperintensities present in 179 (49%) and median modified total SVD score of 1.0 [0.0, 2.0] (Table 1).

On central adjudication (Table 2), a visible index infarct was present in 319 (88%) and averaged 10×8×10 mm in size, Only one scan was considered to be normal, i.e. to have neither an acute infarct responsible for the recent stroke symptoms or any background changes of SVD or atrophy. White matter hyperintensities were present in 95% of participants: 76% had up to Fazekas 2 periventricular and 82% had up to Fazekas 2 deep WMH scores. 62% of participants had one or more old vascular lesions including lacunes present in 96%. Microbleeds were present in 20% of patients in mainly lobar (28%), deep (41%) or mixed lobar/deep (31%) locations. Atrophy was present in 75% of participants.

**Table 2.**
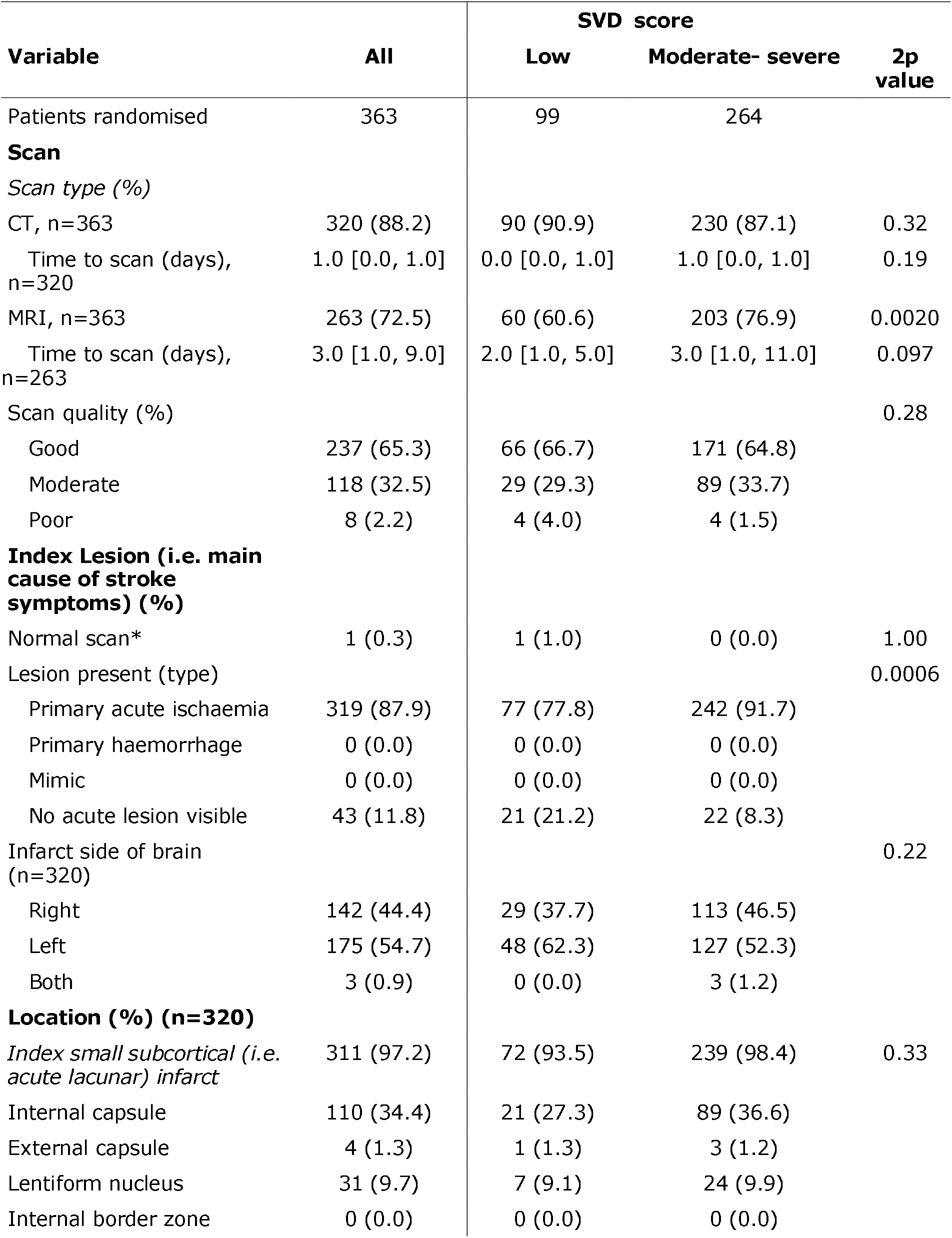
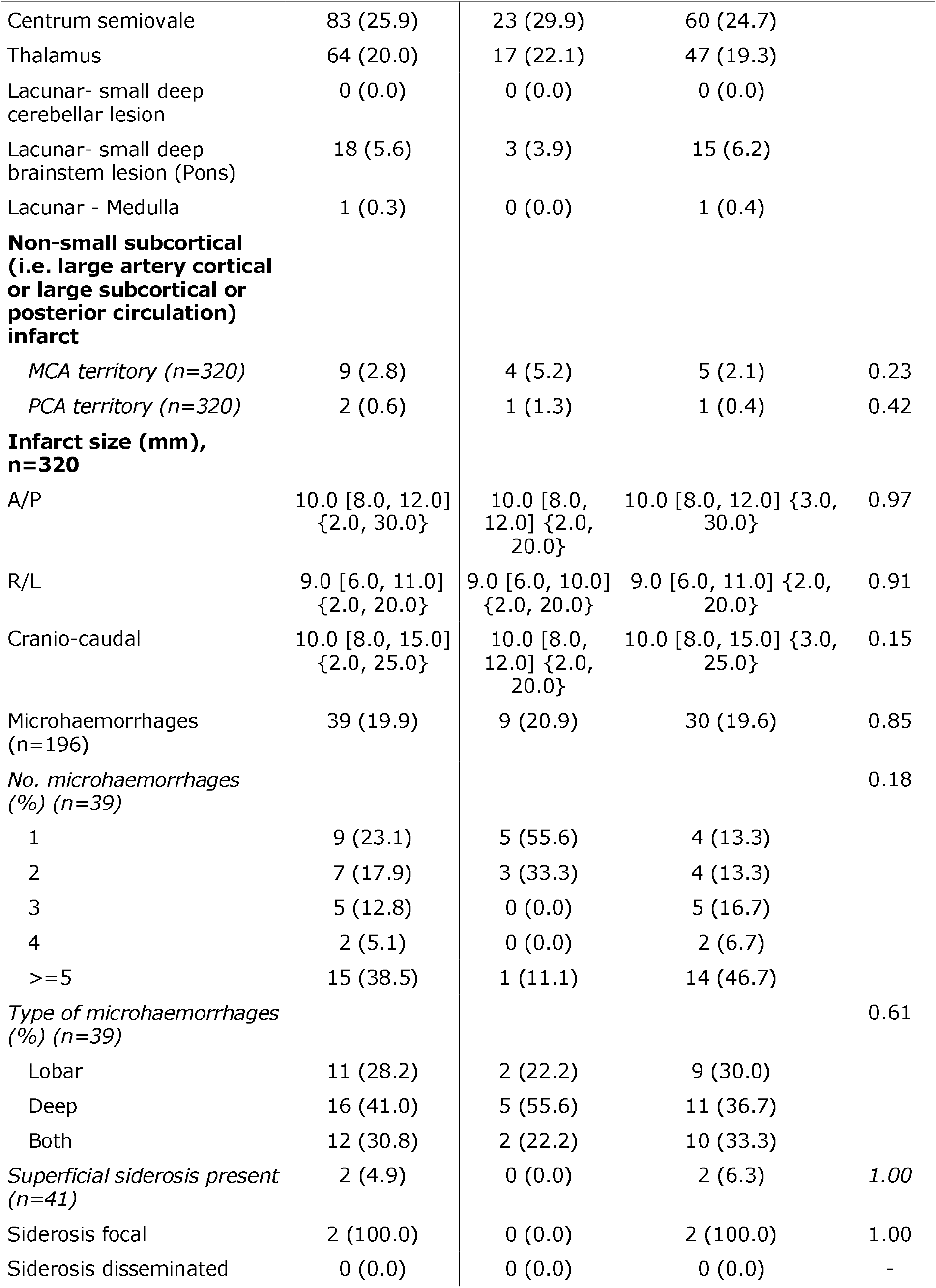
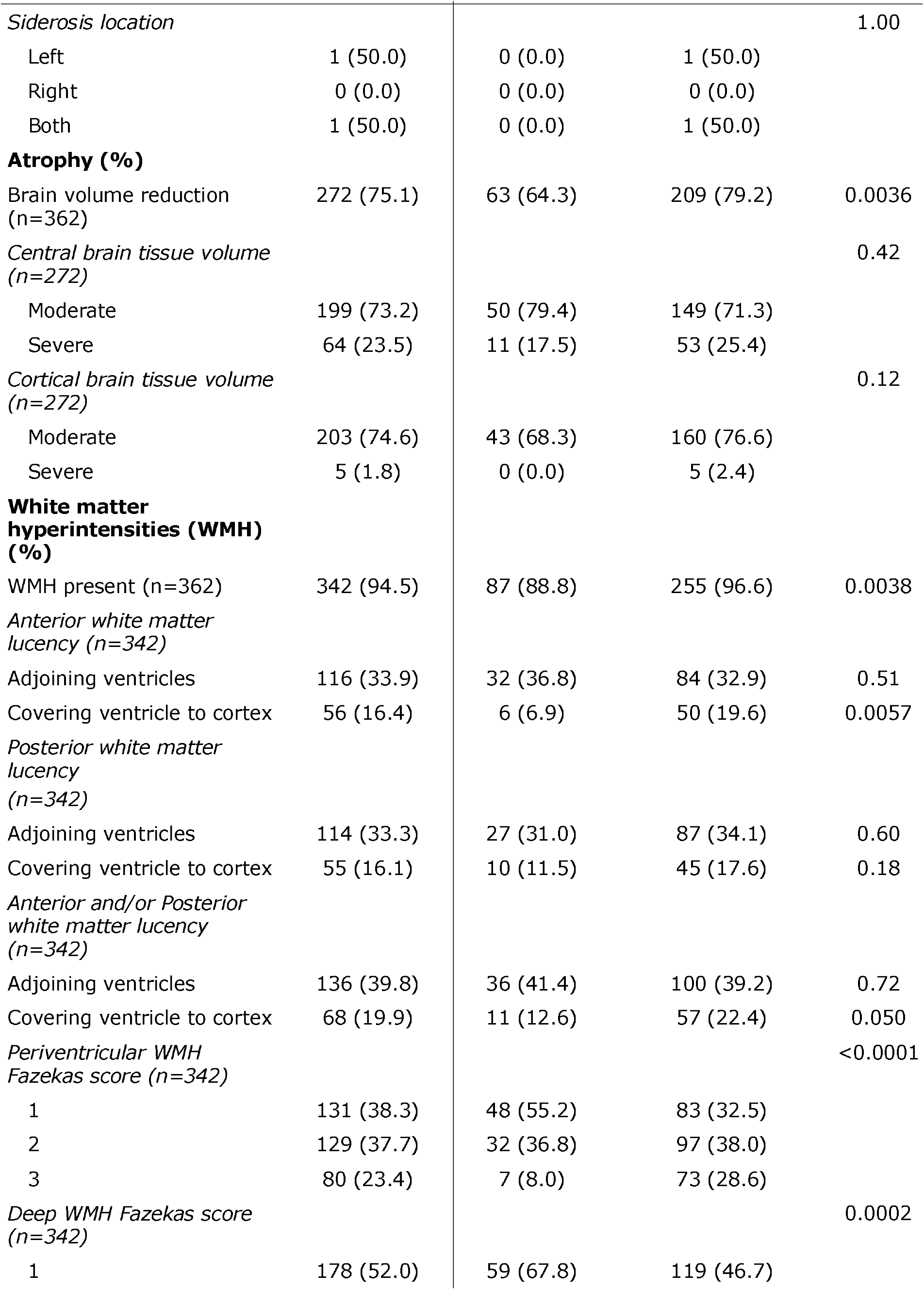
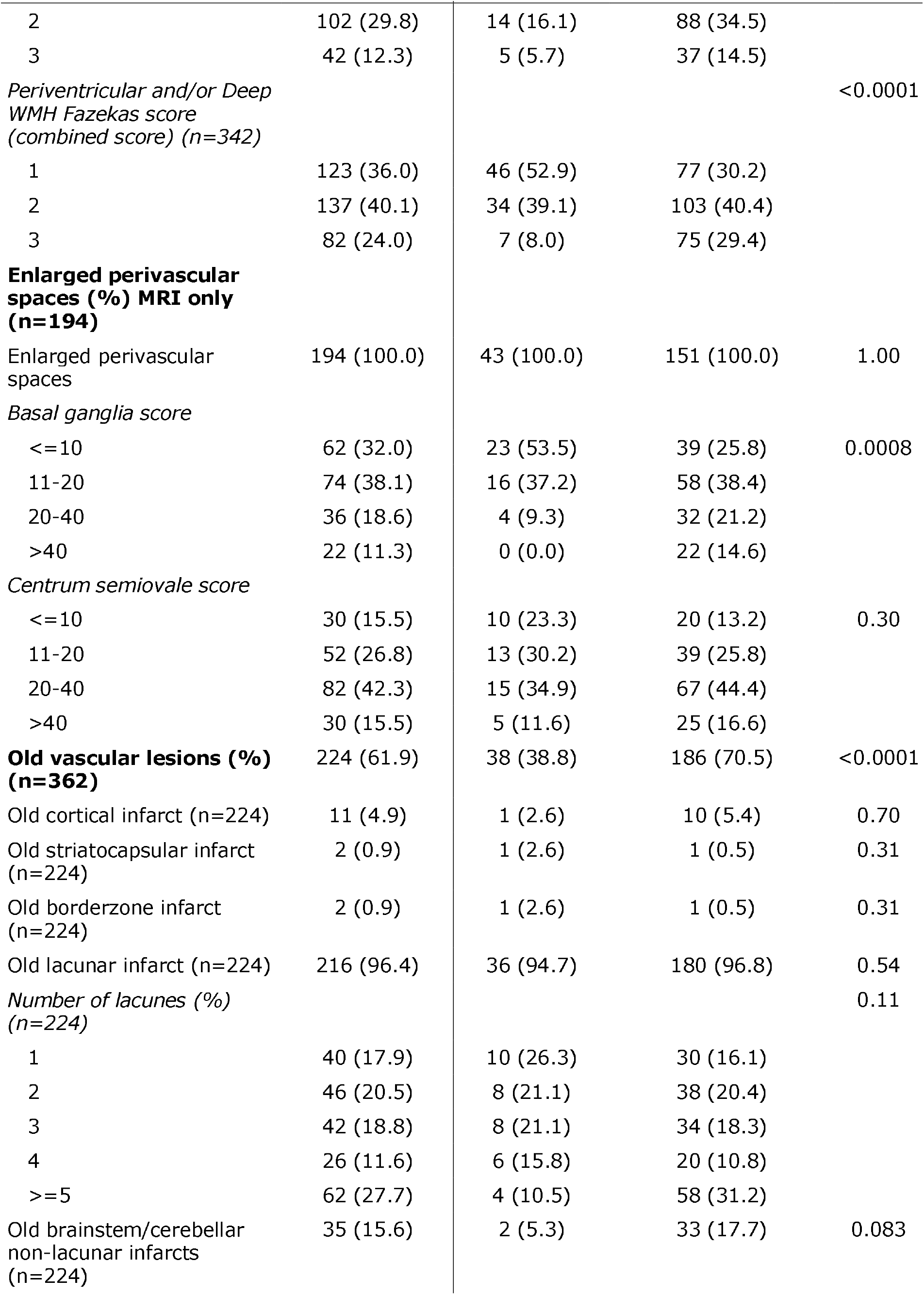
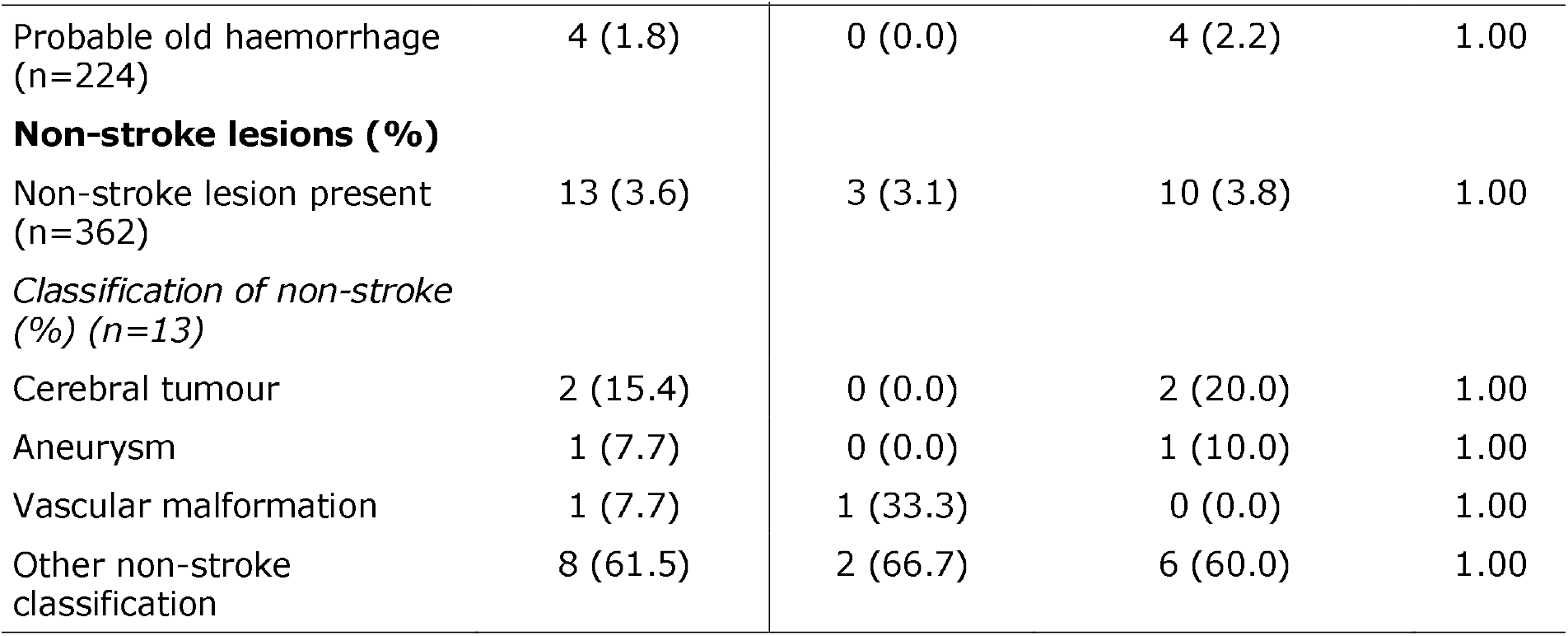
Adjudicated baseline imaging characteristics by small vessel disease score as determined by recruiting site. Data are number (%), median [IQR], or mean (standard deviation). Number of participants with data is 363 unless stated.

In univariate analyses and in comparison with participants with lower SVD scores (Table 1), those with moderate or severe SVD scores were older and more likely to have had a previous stroke, be taking antihypertensive drugs and have ataxia, and less likely to have visual loss. Additionally, participants with more severe SVD scores were more likely to have had MRI at diagnosis and to have a relevant acute ischaemic stroke lesion present on imaging. Similarly, in comparison with participants with a low SVD score, those with a moderate/high score had more atrophy, white matter hyperintensities and their severity, and the presence of old vascular lesions (Table 2).

## DISCUSSION

Small vessel disease is common world-wide and, so far, whether presenting covertly or with stroke, cognitive or physical impairment, or neuropsychiatric symptoms,^28^ has no demonstrated interventions that prevent or limit its development and progression. ^2-4 16^ The LACI trials are assessing the tolerability and feasibility of recruiting patients, the outcome event rates, and tolerability and safety of treatment with ISMN and cilostazol ^17 21-24^ in addition to guideline secondary stroke prevention, with the aim of testing these agents in a large phase III efficacy trial.

The baseline characteristics presented here show that LACI-2 is representative of patients with clinically-evident lacunar ischaemic stroke, a marker of SVD. In particular, the average age was younger than for other types of ischaemic stroke at 64 years,^29^ there was a preponderance of males (69%) ^30^ and most participants had relatively few physical signs or impairments resulting from their stroke.^15^ A majority of participants were diagnosed with hypertension and taking antihypertensive medication, and most were taking antiplatelet or lipid lowering drugs as guideline secondary prevention of ischaemic stroke. Many were ex- or current smokers. Very few had atrial fibrillation or carotid stenosis (about 1%) reflecting that, while emboli can enter and intracranial large artery atheroma may obstruct perforating cerebral arterioles, these are rare in, and uncommon causes of, lacunar ischaemic stroke.^29 31^

A minority of participants had clinical cortical features such as dysphasia (19, 5.2%), neglect (6, 2%), or hemianopia (11, 3%), reflecting the overlap in symptoms between cortical and lacunar ischaemic stroke,^18^ and potential difficulty of recruiting pure lacunar syndrome populations on the basis of clinical syndrome and CT scanning in busy regional hospital stroke services. Nonetheless, it is reassuring that very few patients were found to have a primary cortical infarct on central expert scan read (3%), the remaining 97% having either a recent small subcortical (i.e. acute lacunar) infarct as the primary cause of symptoms, or no definite visible relevant lacunar infarct but no alternative cause for their symptoms.

On adjudicated brain imaging, most participants (264, 73%) were judged to have a moderate to severe SVD score by the recruiting hospital, consistent with patients with lacunar ischaemic stroke, and demonstrating that a simplified version of the SVD score, suitable for use on CT or MRI, can be applied in busy stroke services.^32^

The trial is in follow-up and once this is completed, the database will be cleaned and locked. Therefore there may be minor changes in the baseline data between that provided here and in subsequent publications. Analysis will follow the statistical analysis plan given here as a supplement. If reasonable drug adherence and safety are confirmed, then a phase III trial will be designed and submitted for funding using the vascular and cognitive outcome event rates to power the next phase of the trial.

## Supporting information

Statistical analysis plan

## Data Availability

The baseline data form part of the main trial database and will be used for covariate adjustment. The individual patient baseline data are not available although the whole trial database will become available eventually.

## TRIAL REGISTRATION

The trial is registered: EudraCT 2016–002277-35, ISRCTN 14911850.

## ACKNOWLEDGEMENTS

We thank the patients and their relatives for their time and effort to participate in LACI-2; and the Trial Steering Committee, Sponsor, Data Monitoring Committee, Edinburgh Clinical Trials Unit staff, UK Clinical Research Network, Scottish Stroke Research Network, International Advisory Panel, and all staff at participating sites (listed in the protocol paper ^17^), for their support.

## ETHICAL APPROVAL

The trial is conducted in accordance with the principles of the International Conference on Harmonisation Tripartite Guideline for Good Clinical Practice (ICH GCP). Ethics approval was granted by the East Midlands Nottingham 2 Research Ethics Committee of the Health Research Authority number 17/EM/0077 on 10/05/2017. NHS Research and Development Approval is given by each participating Centre. The Medicines and Healthcare Regulatory Authority approved the trial on 01/06/2017. LACI-2 was adopted by the UK Clinical Research Network and Scottish Stroke Research Network.

## INFORMED CONSENT

Participants must have capacity and consent is taken from them for the trial and for secondary data uses; consent is taken from a relative or other informant to provide outcome data.

## CONTRIBUTIONS

JMW, PMB, FD, NS, VC, THE, AH, DW designed the trial and secured funding; JMW is the Chief Investigator, obtained ethics and regulatory approvals; PMB is Director of the Nottingham Stroke Trials Unit. GB, JPA helped design the assessments, and with JMW and PMB, designed the case record form; KO is the trial manager, responsible for daily running of the trial including regulatory compliance; FD, THE, AH, DW are Principle Investigators; NS and FD provide independent blinded event review; AM provides statistical expertise; IM and LW are the trial statisticians; PMB drafted the manuscript; all other authors commented and edited it; all authors approved the final version for submission.

## REFERENCES

1. Wardlaw JM, Smith C, Dichgans M. Small vessel disease: mechanisms and clinical implications. Lancet Neurol 2019 doi: 10.1016/S1474-4422(19)30079-1 [published Online First: 2019/05/13]

2. Bath PM, Wardlaw JM. Pharmacological treatment and prevention of cerebral small vessel disease: a review of potential interventions. Int J Stroke 2015;10(4):469–78. doi: 10.1111/ijs.12466

3. SPS3 Investigators BO, Hart RG, McClure LA, Szychowski JM, Coffey CS, Pearce LA. Effects of clopidogrel added to asprin in patients with recent lacunar stroke. New England Journal of Medicine 2012;376(9):817–25. doi: 10.1056/NEJMoa1204133

4. Benavente OR, Coffey CS, Conwit R, et al. Blood-pressure targets in patients with recent lacunar stroke: the SPS3 randomised trial. Lancet 2013;382(9891):507–15. doi: 10.1016/s0140-6736(13)60852-1 [published Online First: 29 May 2013]

5. Appleton J, Scutt P, Sprigg N, et al. Hypercholesterolaemia and vascular dementia. Clinical Sciences 2017;131(14):1561–78. doi: 10.1042/CS20160382 [published Online First: 21 March 2017]

6. Bath PMW, Hassall DG, Gladwin A-M, et al. Nitric oxide and prostacyclin. Divergence of inhibitory effects on monocyte chemotaxis and adhesion to endothelium in vitro. Arteriosclerosis and Thrombosis 1991;11:254–60.

7. Leonardi-Bee J, Bath PM, Bousser MG, et al. Dipyridamole for preventing recurrent ischemic stroke and other vascular events: a meta-analysis of individual patient data from randomized controlled trials. Stroke 2005;36(1):162–8. doi: 10.1161/01.STR.0000149621.95215.ea [published Online First: 2004/12/01]

8. McHutchison C, Blair GW, Appleton JP, et al. Cilostazol for Secondary Prevention of Stroke and Cognitive Decline: Systematic Review and Meta-Analysis. Stroke 2020;51(8):2374–85. doi: 10.1161/STROKEAHA.120.029454 [published Online First: 20200710]

9. Rajani RM, Quick S, Ruigrok SR, et al. Reversal of endothelial dysfunction reduces white matter vulnerability in cerebral small vessel disease in rats. Sci Transl Med 2018;10(448) doi: 10.1126/scitranslmed.aam9507

10. Hasel P, Dando O, Jiwaji Z, et al. Neurons and neuronal activity control gene expression in astrocytes to regulate their development and metabolism. Nat Commun 2017;8:15132. doi: 10.1038/ncomms15132 [published Online First: 20170502]

11. Sprigg N, Gray LJ, Bath PMW, et al. Stroke severity, early recovery and outcome are each related with clinical classification of stroke: Data from the ‘Tinzaparin in Acute Ischaemic Stroke Trial’ (TAIST). Journal of the Neurological Sciences 2007;254(1-2):54–59. doi: Doi 10.1016/J.Jns.2006.12.016

12. Sprigg N, Gray LJ, Bath PM, et al. Early recovery and functional outcome are related with causal stroke subtype: data from the tinzaparin in acute ischemic stroke trial. J Stroke Cerebrovasc Dis 2007;16(4):180–4. doi: 10.1016/j.jstrokecerebrovasdis.2007.02.003 [published Online First: 2007/08/11]

13. Georgakis MK, Duering M, Wardlaw JM, et al. WMH and long-term outcomes in ischemic stroke: A systematic review and meta-analysis. Neurology 2019;92(12):e1298–e308. doi: 10.1212/WNL.0000000000007142 [published Online First: 20190215]

14. Debette S, Schilling S, Duperron MG, et al. Clinical Significance of Magnetic Resonance Imaging Markers of Vascular Brain Injury: A Systematic Review and Meta-analysis. JAMA Neurol 2019;76(1):81–94. doi: 10.1001/jamaneurol.2018.3122

15. McHutchison CA, Cvoro V, Makin S, et al. Functional, cognitive and physical outcomes 3 years after minor lacunar or cortical ischaemic stroke. J Neurol Neurosurg Psychiatry 2019;90(4):436–43. doi: 10.1136/jnnp-2018-319134 [published Online First: 20181215]

16. Wardlaw JM, Debette S, Jokinen H, et al. ESO Guideline on covert cerebral small vessel disease. Eur Stroke J 2021;6(2):IV. doi: 10.1177/23969873211027002 [published Online First: 20210618]

17. Wardlaw J, Bath PMW, Doubal F, et al. Protocol: The Lacunar Intervention Trial 2 (LACI-2). A trial of two repurposed licenced drugs to prevent progression of cerebral small vessel disease. Eur Stroke J 2020;5(3):297–308. doi: 10.1177/2396987320920110 [published Online First: 20200420]

18. Potter G, Doubal F, Jackson C, et al. Associations of clinical stroke misclassification (‘clinical-imaging dissociation’) in acute ischemic stroke. Cerebrovascular diseases (Basel, Switzerland) 2010;29(4):395–402. doi: 10.1159/000286342 [published Online First: 2010/02/23]

19. Makin SD, Doubal FN, Dennis MS, et al. Clinically Confirmed Stroke With Negative Diffusion-Weighted Imaging Magnetic Resonance Imaging: Longitudinal Study of Clinical Outcomes, Stroke Recurrence, and Systematic Review. Stroke 2015;46(11):3142–8. doi: 10.1161/STROKEAHA.115.010665 [published Online First: 20150929]

20. Appleton JP, Woodhouse LJ, Adami A, et al. Imaging markers of small vessel disease and brain frailty, and outcomes in acute stroke. Neurology 2020;94(5):e439–e52. doi: 10.1212/WNL.0000000000008881 [published Online First: 2019/12/27]

21. Blair GW, Appleton JP, Law ZK, et al. Preventing cognitive decline and dementia from cerebral small vessel disease: The LACI-1 Trial. Protocol and statistical analysis plan of a phase IIa dose escalation trial testing tolerability, safety and effect on intermediary endpoints of isosorbide mononitrate and cilostazol, separately and in combination. Int J Stroke 2017:1747493017731947. doi: 10.1177/1747493017731947 [published Online First: 2017/09/15]

22. Appleton JP, Blair GW, Flaherty K, et al. Effects of Isosorbide Mononitrate and/or Cilostazol on Hematological Markers, Platelet Function, and Hemodynamics in Patients With Lacunar Ischaemic Stroke: Safety Data From the Lacunar Intervention-1 (LACI-1) Trial. Front Neurol 2019;10:723. doi: 10.3389/fneur.2019.00723 [published Online First: 2019/07/03]

23. Blair GW, Appleton JP, Flaherty K, et al. Tolerability, safety and intermediary pharmacological effects of cilostazol and isosorbide mononitrate, alone and combined, in patients with lacunar ischaemic stroke: The LACunar Intervention-1 (LACI-1) trial, a randomised clinical trial. EClinicalMedicine 2019;11:34–43. doi: 10.1016/j.eclinm.2019.04.001 [published Online First: 2019/04/24]

24. Blair GW, Janssen E, Stringer MS, et al. Effects of Cilostazol and Isosorbide Mononitrate on Cerebral Hemodynamics in the LACI-1 Randomized Controlled Trial. Stroke 2022;53(1):29–33. doi: 10.1161/STROKEAHA.121.034866 [published Online First: 20211201]

25. Gamble C, Krishan A, Stocken D, et al. Guidelines for the Content of Statistical Analysis Plans in Clinical Trials. JAMA 2017;318(23):2337–43. doi: 10.1001/jama.2017.18556

26. Hemming K, Kearney A, Gamble C, et al. Prospective reporting of statistical analysis plans for randomised controlled trials. Trials 2020;21(1):898. doi: 10.1186/s13063-020-04828-8 [published Online First: 20201028]

27. MacMahon S, Collins R. Reliable assessment of the effects of treatment on mortality and major morbidity, II: observational studies. Lancet 2001;357(9254):455–62. doi: 10.1016/s0140-6736(00)04017-4 [published Online First: 2001/03/29]

28. Clancy U, Gilmartin D, Jochems ACC, et al. Neuropsychiatric symptoms associated with cerebral small vessel disease: a systematic review and meta-analysis. Lancet Psychiatry 2021;8(3):225–36. doi: 10.1016/S2215-0366(20)30431-4 [published Online First: 20210201]

29. Jackson CA, Hutchison A, Dennis MS, et al. Differing risk factor profiles of ischemic stroke subtypes: evidence for a distinct lacunar arteriopathy? Stroke 2010;41(4):624–9. doi: 10.1161/strokeaha.109.558809 [published Online First: 2010/02/13]

30. Jiménez-Sánchez L, Hamilton OKL, Clancy U, et al. Sex Differences in Cerebral Small Vessel Disease: A Systematic Review and Meta-Analysis. Front Neurol 2021;12:756887. doi: 10.3389/fneur.2021.756887 [published Online First: 20211028]

31. Del Bene A, Makin SD, Doubal FN, et al. Variation in risk factors for recent small subcortical infarcts with infarct size, shape, and location. Stroke 2013;44(11):3000–6. doi: 10.1161/STROKEAHA.113.002227 [published Online First: 20130905]

32. Staals J, Makin S, Doubal F, et al. Stroke subtype, vascular risk factors, and total MRI brain small-vessel disease burden. Neurology 2014;83:1228–34. doi: 10.1212/WNL.0000000000000837 [published Online First: 27 August 2014]

