## Supplementary material for "Cilostazol and isosorbide mononitrate for the prevention of progression of cerebral small vessel disease: Baseline data and statistical analysis plan for the Lacunar Intervention Trial-2 (LACI-2) (ISRCTN14911850)": Statistical analysis plan

#### SECTION 1. ADMINISTRATIVE INFORMATION

##### 1 Title and trial registration
***1a Title***: Lacunar Intervention Trial 2
***Acronym***: LACI-2
***1b Registration***: ISRCTN14911850; IRAS project number: 206480

**2 SAP version**: 1.5 (06 June 2022)

**3 Protocol version**: 7.0 (14 October 2020)

##### 4 SAP revisions
***4a Revision history***:
Version 1.4 to 1.5
- Text added about the planned soft database lock and analysis (section 13a).
- Text added explaining comparison of dual versus no treatment (section 27a).
- Differences and p values removed (Tables 1, 5).
- Analyses comparing dual versus no treatment do not include all four groups so cilostazol only and ISMN groups removed (Table 9).

***4b Justification for each revision***: N/A
***4c Timing of SAP revisions***: These antedate data lock and analysis. Where there is a difference between the protocol (on website), published protocol [1] and SAP, the SAP will take precedence.

##### 5 Roles and responsibilities
***Author***: Philip M Bath
***Responsible statisticians***: Iris Mhlanga (blinded statistician), Lisa J Woodhouse (blinded statistician), Alan A Montgomery
***Chief Investigator***: Joanna M Wardlaw
***Contributors and roles***:
Philip M Bath, Iris Mhlanga, Lisa J Woodhouse, Fergus Doubal, Katherine Oatey, Alan A Montgomery, Joanna M Wardlaw, for the LACI-2 Investigators*

##### 6 Signatures

| Role | Name | Signature | Date |
|---|---|---|---|
| ***6a Author***: | Philip Bath | Philip Bath<br>Philip Bath (Jun 9, 2022 10:02 GMT+1) | Jun 9, 2022 |
| ***6b Senior statistician*** | Alan Montgomery | A. Montgomery | Jun 15, 2022 |
| ***6c Chief Investigator***: | Joanna Wardlaw | Joanna Wardlaw | Jun 9, 2022 |

#### SECTION 2. INTRODUCTION

##### 7 Background and rationale

Prior to analysis and presentation of the primary results, this publication presents the statistical analysis plan (SAP) [2 3] alongside the detailed listing of baseline characteristics presented in the accompanying baseline paper. This Supporting Information Appendix S1 details the full SAP and is presented prior to locking of the study database so that analyses are not data driven or reported selectively.[4] In addition to the SAP, we also list planned secondary analyses and substudies. The SAP follows the recommended layout.[2 3]

**8 Objectives**

***8a Primary Objective***: To determine whether a prospective, randomised trial of cilostazol and ISMN, individually or in combination, on a background of guideline stroke prevention therapy, in lacunar ischaemic stroke is feasible in the UK, thence proceeding as seamlessly as possible to a large phase III trial.

***8b Secondary Objectives***: To assess drug tolerability, safety, recruitment rates and accuracy, outcome event rates and retention in preparation to a large phase III randomised controlled trial to prevent recurrent lacunar stroke and physical and cognitive impairment.

This SAP focuses on these primary and secondary objectives. Planned follow-on publications will address tertiary questions.

#### SECTION 3. STUDY METHODS

**9 Trial design**

Prospective randomised open-label blinded end-point (PROBE) partial-factorial phase IIb/c trial aiming to recruit 400 patients recruited in UK Stroke Network Centres, with follow-up to one year.

**10 Randomisation**: By central computer-generated allocation at the University of Nottingham with minimisation on key prognostic factors: age, sex, stroke severity (NIHSS), dependency resulting from the stroke, systolic blood pressure ≤/>140 mmHg, smoking status, time after stroke, and years of education.

**11 Sample size/power considerations**

Conservatively, we have used sample size calculations based on binary measures. Use of ordinal measures at the time of analysis will increase statistical power.

***11a Event rates***: Annual event rates (Table A) were assessed from trials (SPS3,[5] lacunar patients in ENOS,[6 7] IST-3 [8 9]) and observational data (LADIS;[10] our [11-13] and other [14] studies). All-cause death rates were assumed to be 2.0% with upper 95% CI of 4% in 400 patients.[5] Hence, the sample size was set at 400 participants.

**Table A** Annual absolute risks (%) of outcome events after lacunar stroke

| Vascular death | Non-vascular death | Non-fatal IS or TIA | Non-fatal ICH | MI | MACE | Dependent (mRS 3-5) | Cognitive impairment | Dementia |
|---|---|---|---|---|---|---|---|---|
| 1.8 | 0.5 | 2.5 | 0.5 | 0.6 | 3 | 15 | 30 | 15 |

ICH: intracerebral haemorrhage; IS: ischaemic stroke; MACE: major adverse cardiac events; MI: myocardial infarction; mRS: modified Rankin scale; TIA: transient ischaemic stroke

***11b Comparison of two groups in a future phase III trial***: Assuming power 0.80, alpha=0.05, 1:1 randomisation, composite event rate (MACE, dementia, non-vascular death, new MRI signs) 45% and absolute reduction 9% (relative risk reduction 20%), and loss to follow-up 10%, a sample size of 1100 will be needed. A number of outcomes are relevant to patients with SVD and using these has implications for the sample size (Table B).

**Table B.** Sample size for composite outcome in main trial using estimated event rates.[1]

| Composite model | A | B | Ci | CiI | D |
|---|---|---|---|---|---|
| Composite outcome for phase III | MACE, dementia, non-vascular death, new MR signs | MACE, dementia, death | MACE, cognitive decline, dependency decline, all-cause death | | MACE, cognitive impairment, dependency, all-cause death |
| 1-beta (power) | 80% | 80% | 80% | 80% | 80% |
| Event rate, control, pa | 50% | 10% | 30% | 30% | 45% |
| Relative risk reduction | 20% | 20% | 20% | 30% | 20% |
| Event rate, active, pa | 40% | 8% | 24% | 21% | 36% |
| Total sample size | 950 | 6626 | 1784 | 778 | 976 |
| Total trial size, including losses | 1250 | 7400 | 2000 | 900 | 1100 |

MACE: major adverse cardiac events; MRI: magnetic resonance imaging

#### 12 Framework

The primary objectives are to assess the feasibility of recruitment and adherence to medication.

#### 13 Statistical interim analyses and stopping guidance

***13a Interim analyses***: Data are tabulated twice annually prior to Trial Steering Committee (TSC) and Data Monitoring Committee (DMC) meetings. No unblinded comparative analyses will be performed until data collection has been completed and the database locked.

Prior to the final database lock, the database will be subject to a soft lock and the provisional data tabulated and analysed for review by the TSC and DMC. Any final queries will be raised and resolved prior to final database lock. Members of staff still involved in the collation of data and resolution of data queries will not attend the meeting and the data reviewed at the meeting will remain strictly confidential until the point of final database lock to avoid any bias.

***13b Adjustments of significance level***: There is no planned adjustment.
***13c Stopping rules***: There are no formal stopping rules, but the DMC have responsibility to make recommendations to pause or modify the study, should there be any safety or efficacy considerations.

#### 14 Timing of final analyses

These will be performed once data collection has been completed and the database has been locked.

#### 15 Timing of outcome assessments

Assessments will be performed at baseline, 1-2 weeks, 3-4 weeks, 6 and 12 months (***Table C***).

**Table C**. Assessments at baseline and follow-up by time point (adapted from protocol and [1]).

| Assessment | Prior to Baseline | Visit 1 Baseline | Week 1-2 | Week 3-4 | Month 6 | Month 12 |
|---|---|---|---|---|---|---|
| Screening for eligibility and consent [†] | X[S] | | | | | |
| Confirm and document ongoing consent | | X[S] | | | | |
| Medical including drug history | | X[S] | | | | |
| Assess MR or CT diagnostic scan; send copy to Edinburgh | | X[S] | | | | |
| Randomisation | | X[S] | | | | |
| Haematology (full blood count) and Biochemistry (urea, electrolytes, creatinine) – most recent value obtained since time of index stroke is acceptable unless clinical reason to expect change | | X[S] | | | | |
| Blood pressure | | X[S] | | | | X[S ‡] |
| Cognitive test: document years of education; Montreal Cognitive Assessment (MOCA) | | X[S] | | | | |
| Timed Trail Making Test B | | X[S] | | | | X[S ‡] |
| Dispense trial medication[2] | | X[S] | | | X[S] | |
| Structured questionnaire: symptoms; medication history and IMP tablet adherence | | | X[S] | X[S] | X[C] | X[C] |
| Structured questionnaire: recurrent vascular events, mRS, TICS, t-MOCA, SIS, ZUNG | | | | | X[C] | X[C] |
| Obtain IQCODE (post/phone) from relative | | | | | | X[C] |
| Follow-up brain MRI | | | | | | X[S] |
| Health Economics data: EQ-5D-5L, EQ-VAS | | | | | | X[C] |
| Adverse event / con meds reporting as necessary | | | X[S] | X[S] | X[S,C] | X[S,C] |

[†] Consent will be obtained before the data collection procedures commence or randomisation is performed. Randomisation occurs at the end of the baseline visit.

[‡] at 12 months in some centres only.

[2] Dispensing in 3-monthly intervals is allowed.

[S] Assessment performed by local site team.

[C] Assessment performed by blinded assessor who is part of the central trial team.

SIS: Stroke Impact Scale; TICS: telephone interview for cognitive status; t-MOCA: telephone MOCA.

#### SECTION 4. STATISTICAL PRINCIPLES

**Confidence and p values**

**16 Level of statistical significance**

The results of analyses and comparisons will be shown with p<0.05.

**17 Multiplicity**

No adjustment will be made for multiplicity.

**18 Levels of confidence intervals**

The results of analyses and comparisons will be shown with 95% confidence intervals.

**19 Adherence**

***19a Definition***: 75% of patients will be able to tolerate trial medication, in at least half dose, up to one year after randomisation (i.e. less than 25% will stop trial medication completely through inability to tolerate the drugs).
***19b Adherence presentation***: See Table 4.
**19c Protocol deviations**: Protocol violations will be reported to the sponsor within 24 hours of becoming aware of the violation. Protocol deviations will be recorded in a protocol deviation log with these submitted to the sponsors every 3 months.
***19d Protocol deviation presentation***: Listing of violations and deviations and their frequency.

**20 Analysis populations**

Three populations are defined:

1. Intention-to-treat: All consented participants with a primary outcome measure.
2. Per protocol: All consented participants with a primary outcome measure who received at least one dose of randomised medication and who had no protocol violation, e.g. they fulfilled all eligibility criteria.
3. Safety: All consented participants who received at least one dose of randomised medication.

Multiple variable analyses will include all patients with complete data for the dependent and each independent variable. All available data will be used, and missing data will not be imputed.

#### <u>SECTION 5. TRIAL POPULATION</u>

**21 Screening data**

No screening logs will be kept so that data collection can be prioritised.

**22 Eligibility**

1. Clinical lacunar stroke syndrome.
2. Brain scanning with MR including diffusion imaging wherever possible, and obtained soon after the presentation with stroke, which showed either:
    a. A recent, relevant (in time and location) acute small subcortical (i.e. lacunar) infarct on diffusion MR imaging.
    b. If no visible acute small subcortical infarct on diffusion MR imaging then there is no competing pathology as a cause for stroke (e.g. no acute cortical infarct, no acute intra-cerebral haemorrhage, no stroke mimic such as tumour, subdural haematoma);
    c. If only a CT brain scan is available, then there is a small relevant (in time and location) subcortical (i.e. acute lacunar) infarct, or if no infarct then there is no competing pathology as a cause for stroke (e.g. no acute cortical infarct, no acute intra-cerebral haemorrhage, no stroke mimic such as tumour, subdural haematoma).

3. Age >30 years.
4. Independent in activities of daily living (modified Rankin Scale <=2).
5. Capacity to give consent themselves.

**23 Recruitment**
Recruitment will be summarised in a CONSORT flow diagram.

**24 Withdrawal/follow-up**
Withdrawals, and missed follow-ups and their timing will be summarised in the CONSORT flow diagram.

**25 Baseline patient characteristics**
***25a Baseline characteristics***: These will comprise demographic, education, premorbid function, cognitive ability, medical history, blood pressure, stroke investigations, clinical and brain imaging parameters (Table 1).
***25b Summarisation***: Data will be shown as number (%), median [interquartile range] or mean (standard deviation) as appropriate.

#### SECTION 6. ANALYSIS

**26 Outcome definitions**
***26a Specifications:***
***Primary endpoint***
Feasibility of a Phase III efficacy trial assessed as:
- Recruitment of sufficient patients, i.e. 400 patients in 24 months in the UK (and taking account of interruption due to COVID-19).
- >95% of randomised patients are retained for follow-up at one year.

***Secondary outcomes - participant***
*Tolerability*: 75% of patients will be able to tolerate trial medication, in at least half dose, up to one year after randomisation (i.e. less than 25% will stop trial medication completely through inability to tolerate the drugs).
*Safety*
- Symptoms of systemic or intracranial bleeding.
- The absolute risk of death, including fatal haemorrhage, does not differ significantly, i.e. fall outside the upper 95% CI of 2% per year on trial drugs versus no trial drugs, when given in addition to guideline stroke prevention drugs.
- There are no new ischaemic or haemorrhagic brain lesions or increase in SVD lesions on one year MRI significantly (at the p<0.01 level).

*Efficacy*
- Individual event-rates for stroke, TIA, myocardial ischaemia, cognitive impairment and dementia.
- The *combined rate* of recurrent stroke, MI, death, mild cognitive impairment (including dementia), dependency and new stroke lesions on scanning at 1 year will be 40-50% at one year after enrolment in order to allow detection of a modest but clinically-important reduction in poor outcomes in a phase III trial.
- Health economic measures include the health utility score (EQ-5D-5L) and the visual analogue score (EQVAS) at 12 months.

***26b Units***: Units will be shown in tables.
***26c Calculations/transformations***: Quality of life using UK weightings.
***Brain frailty***
Based on neuroimaging:
- Brain frailty = Atrophy + WML + Previous stroke lesion [7]

- SVD score for CT = WML, lacunes; for MRI includes WMH, lacunes, PVS and microbleeds [15]

***Montreal cognitive assessment-modified (MoCA-m) trails***

Since Trails A and B are performed, the MoCA trail is not collected but rather estimated from the Trails B score:

- If Trails B score <12 then MoCA trail = 0
- If Trails B score >=12 then MoCA trail = 1

#### 27 Analysis methods

##### 27a Methods

*Primary endpoint*

- Tabulation and graphical presentation of participant recruitment aiming for 400 participants in 24 months.
- Tabulation of retention of participants at one year aiming for >95%.

*Analyses of secondary outcomes*

Tabulations of:

- Tolerability to trial medications aiming for 75% of patients taking at least half dose for up to one year after randomisation.
- Death, including fatal haemorrhage, aiming for less than outside the upper 95% CI of 2% per year.
- The *combined rate* of recurrent stroke, MI, death, cognitive impairment and dependency, aiming for 40-50% at one year after enrolment.

Comparison of rates of events between the treatment groups: cilostazol vs no cilostazol, ISMN vs no ISMN, and cilostazol and ISMN vs neither, for:

- Systemic or intracranial bleeding, recurrent cerebral and systemic vascular events, and vascular and non-vascular causes of death.
- Death, all cause.
- New ischaemic or haemorrhagic stroke lesion or increase in SVD lesions on MRI.
- Composite of: recurrent clinically-evident stroke, MI, death, cognitive impairment (including dementia) and dependency.
- Individual event: stroke (clinically-evident or imaging-detected ischaemic or haemorrhagic stroke to be reported separately), TIA, myocardial ischaemia, cognitive impairment and dementia.
- New infarcts and haemorrhages, absolute and change in WMH, microhaemorrhages, lacunes, atrophy imaging variables from central read of baseline imaging and one year MRI
- The comparisons of combined cilostazol and ISMN versus neither, whilst being very underpowered statistically, are presented since these may be the two groups studied in the planned follow-on trial.

Central tendency, comparisons and regressions will be analysed as follows (***Table D***).

**Table D. Descriptive and analytical statistics**

| | Binary | Nominal | Ordinal | Continuous |
|---|---|---|---|---|
| Central tendency and distribution | N (%) | N (%) | Median [interquartile range] | Mean (standard deviation) |
| Comparisons | Chi-square (2x2) | Chi-square (2x2, or rxc) | Mann-Whitney U or Kruskal-Wallis | t-test (pooled) or 1-way ANOVA |
| Regression | Binary logistic regression (BLR) | - | Ordinal logistic regression (OLR) | Multiple linear regression (MLR) |

***27b Covariate adjustment***: Analyses will be adjusted for minimisation covariates:

- Age, sex, stroke severity (NIHSS), dependency resulting from the stroke, systolic blood pressure, smoking status, time after stroke, years of education.

Covariate adjustment with continuous variables (age, NIHSS, SBP, time after stroke, years of education) will use original, not dichotomised, data.

***27c Assumption checking***: The assumption of proportionality will be tested using the likelihood test.

***27d Alternative methods***: If the data fail the assumption of proportionality (tested using the likelihood test), we will use alternative methods such as multiple logistic regression.

***27e Sensitivity analyses***: In addition to assessment of raw data, the primary outcome will be analysed using additional statistical approaches in sensitivity analyses:

- Unadjusted analysis
- Imputation (multiple regression imputation) of missing data (adjusted) [16]

***27f Subgroup analyses***: The secondary endpoint of the composite of: recurrent stroke, MI, death, cognitive impairment and dependency, will be studied in:

- Pre-specified subgroups comprising the minimisation variables.
- Any other variables demonstrating imbalance at baseline.

The results of these subgroup analyses will not be adjusted for multiple testing. These analyses are planned for the phase III efficacy trial and so will be tested in the present study.

**28 Missing data**

Missing data may occur at outcome level or at test level or within a test at component item level. Notably, some tests have to exclude components if performed by telephone and/or postal questionnaire or require a one year MRI. There is often a relationship between inability to complete outcome assessment and cognitive function or neurological deficit after stroke (e.g. inability to hold a pen) and so assumptions around random missingness may not be valid, even if the patterns of missing data initially suggest 'missing completely at random' status. Indeed, failure to complete a test may be an indicator of cognitive impairment rather than real missingness.

The approaches taken to missing cognitive and other data can have a substantial effect on epidemiological estimates.[16] We will use the approach that makes greatest use of available data.

Where in study data are not available, or participants are lost to follow-up, we have permissions to allow for linkage of the study dataset to primary and secondary care electronic health records. This will allow for an assessment of clinical outcomes across all the participants.

**29 Additional analyses**

***Global outcomes***

We will assess global outcomes integrating multiple scores into one analysis and so provide a more holistic measure and improve statistical power. We will assess global outcomes comprising:

- Recurrent ordinal stroke (clinically-evident and/or imaging-detected),[17] ordinal MI, cognition (MoCA), dependency (mRS), quality of life (EQ-5D).

Analyses will use the Wei-Lachin test [18-21] with comparison of data at 1 year.

***Cognitive domains, based on DSM-V***

We will categorise cognition into 7- and 4-level ordinal scales based on DSM-V [22] categorisation (Table E).[23] We will calculate scores for cognitive domains using sub-scores of MoCA (or TICS if missing) although we recognise that these global cognitive assessments have some test items that map to a more than one domain, e.g. the clock drawing test in the MoCA includes aspects of attention, executive function and visual-perceptual function.

- Learning and memory: orientation in place (from MoCA), delayed recall of five word (MoCA), and recall and delayed recall of ten words (TICS)
- Language: using comprehension, semantic and recent memory (from MoCA; similar elements in TICS-M)
- Perceptual-motor function: Cube copy and clock drawing from MoCA
- Executive function: Trail making tests A & B, verbal fluency test (VFT-phonemic)-F (from MoCA); verbal fluency test (VFT-semantic)-animals; clock drawing test (from MoCA); digits forward (from MoCA); digits backward (from MoCA)
- Complex attention: using serial sevens subtraction (MoCA), letter tapping (MoCA)
- Social cognition is not classically assessed in cognitive screening tools and there are no agreed generic short form assessments for social cognition. Aspects of social cognition will be assessed through informant data and NPI-Q although these are not part of the core outcome set.

**Table E. Categorisation of cognition based on DSM V with operationalisation (adapted summary from [23]).**

<table>
<tr><th></th><th>Seven-level categorisation<br>and operationalisation</th><th>Four-level categorisation and operationalisation</th></tr>
<tr><td>Normal cognition</td><td>No evidence of cognitive impairment<br>(T-MoCA:20-22 AND TICS-m: 25-39)</td><td>No evidence of cognitive impairment<br>(T-MoCA:20-22 AND TICS-m: 25-39)</td></tr>
<tr><td rowspan="2">Minor Neurocognitive disorder<br>(mild cognitive impairment)</td><td>Single domain<br>Scores are reduced by > 1 point in only one cognitive domain of T-MoCA</td><td rowspan="2">Evidence of cognitive impairment<br>(T-MoCA: 15-19 OR TICS-m: 17-24)<br>AND<br>No evidence of functional impairment<br>(mRS <2 OR no change in mRS if pre-stroke mRS >1)</td></tr>
<tr><td>Multi-domain<br>Scores are reduced by > 1 point in more than one cognitive domain of T-MoCA</td></tr>
<tr><td rowspan="3">Major neurocognitive disorder</td><td>Mild cognitive impairments<br>(T-MoCA 15-19 OR TICS-m 17-23)<br>AND<br>Mild dysfunction<br>(mRS <3)</td><td rowspan="3">Persisting cognitive impairment (T-MoCA score <19 OR TICS-m<24 on more than one follow-up)<br>AND<br>Functional impairment<br>(mRS ≥2 or IQCODE >3.6 at final follow-up)<br><br>OR<br><br>Any clinical diagnosis of dementia made independent of study, e.g. by memory clinic, in primary care,<br>recording of dementia on death certification, prescription of cholinesterase inhibitor or memantine</td></tr>
<tr><td>Moderate cognitive impairments<br>(T-MoCA 10-14 OR TICS-m 12-16)<br>AND<br>Moderate dysfunction<br>(mRS 3 or 4)</td></tr>
<tr><td>Severe cognitive impairments<br>(T-MoCA <10 OR TICS-m <12)<br>AND<br>Severe dysfunction</td></tr>
</table>

| | | |
|---|---|---|
| | In a care-home OR mRS 4,5 | |
| **Death** | Death (mRS 6) | Death (mRS 6) |

**30 Harms**
These are presented as serious adverse events in Tables 6a-c and 7a-c.

**31 Statistical software**
Statistical Analysis System (SAS) version 9.4, SAS Institute Incorporation, Cary, North Carolina.

#### SECTION 7. ADDITIONAL INFORMATION

##### Confounding covariates
The primary outcome is recruitment, and this will be tabulated; as such, there are no confounding covariates.

The possible primary outcome in any following trial will likely be a composite comprising MACE, dementia or mild cognitive impairment, non-vascular death and new MRI signs and these event rates will be assessed in the current trial. The components are likely to be correlated. Example confounding variables are given and these are categorised by whether these were 'measured', as per routine practice, or 'unmeasured'; the latter will lead to residual confounding.

##### Composite outcome
**Measured variables**: Age, highest educational attainment, main occupation, socioeconomic status, stroke severity, function at randomisation, cognitive ability at randomisation, diabetes mellitus, hypertension, smoking, carotid disease, blood pressure, prescribed medications, time from stroke to randomisation, presence of a relevant infarct and SVD lesion severity on brain imaging.
***Unmeasured***: Examples are social isolation, vision, hearing, cardiac function (atrial fibrillation and heart failure are documented) and post-stroke complications.

##### Governance
LACI-2 is funded by the British Heart Foundation (CS/15/5/31475) and approved by the East Midlands – Nottingham 2 Research Ethics Committee (Ref: 17/EM/0077). The Sponsor is the ACCORD office, University of Edinburgh and NHS Lothian. NHS Research and Development/ Innovation approval is given at each participating site. The study is adopted by the National Institute for Health Research (NIHR) Clinical Research Network in England and the Stroke Research Network in Scotland.

##### Minimising bias
Multiple approaches are taken to minimise bias: central data registration with real-time on-line validation; minimisation at randomisation; blinded central postal and/or telephone assessment of outcomes; blinded adjudication of neuroimaging; inclusion of patients enrolled in other studies (co-enrolment) where feasible; analysis by intention-to-enrol (i.e. all participants) and in pre-specified subgroups.

##### Publications, published and planned
1. Protocol - published
2. SAP and baseline data - this publication
3. Primary results paper
4. Other secondary publications as determined by the Trial Steering Committee

**Data sharing**

In the future, the anonymised study data will be made available for use by external investigators in appropriate analyses upon request via a publicly accessible portal (e.g. University of Edinburgh data share). Data from LACI-2 will also be shared as appropriate with individual patient data pooling projects involving stroke and dementia; a non-inclusive list includes:

- The Cerebrovascular diseases database, Edinburgh (https://www.ed.ac.uk/clinical-sciences/edinburgh-imaging/research/themes-and-topics/analysis-and-processing/image-databanks/cerebrovascular-diseases-image-databank)
- Dementia Platform UK data portal (https://www.dementiasplatform.uk/)
- Virtual International Stroke Trials Archive-Cognition (VISTA-COG)
- Virtual International Cardiovascular and Cognitive Trials Archive (VICCTA, http://www.virtualtrialsarchives.org)
- META-VCI Map (https://metavcimap.org/)
- STROKOG (https://cheba.unsw.edu.au/consortia/strokog)

Similarly, anonymised neuroimaging data will be published.[24] The mechanisms and processes for managing external access will be determined during the course of the study. Proposals will be considered by the LACI-2 Trial Steering Committee.

#### ABREVIATIONS

| Abbreviation | Full Text |
|---|---|
| CI | Confidence Interval |
| cSVD | Cerebral small vessel disease |
| CT | Computed Tomography |
| DMC | Data Monitoring Committee |
| ECG | Electrocardiogram |
| EQVAS | EuroQol-Visual Analogue Scale |
| EQ-5D-5L | EuroQoL- 5 Dimension- 5 Level quality of life questionnaire |
| F/T | Full Time |
| ICH | Intracerebral Haemorrhage |
| IMP | Investigational Medicinal Product |
| IS | Ischaemic Stroke |
| ISMN | Isosorbide Mononitrate |
| LACI-2 | Lacunar Intervention Trial-2 |
| LACS | lacunar syndrome |
| LVH | left ventricular hypertrophy |
| MACE | Major Adverse Cardiac Events |
| MI | Myocardial Infarction |
| MOCA | Montreal Cognitive Assessment |
| MRI | Magnetic Resonance Imaging |
| mRS | Modified Rankin scale |
| NIHSS | National Institutes Health Stroke Scale |
| NO | Nitric oxide |
| PACS | Partial Anterior Circulation Syndrome; |
| PROBE | Prospective Randomised Open-label Blinded-Endpoint |
| PGI2 | Prostacyclin |
| POCS | Posterior Circulation Syndrome |
| P/T | Part Time |
| SAP | Statistical Analysis Plan |
| SAS | Statistical Analysis System |
| SBP | Systolic Blood Pressure |
| SIS | Stroke Impact Scale |
| SVD | Small vessel disease |
| TACS | Total Anterior Circulation Syndrome |
| TIA | Transient Ischaemic Attack |
| TICS | Telephone Interview for Cognitive Status |
| t-MOCA | Telephone MOCA |
| TSC | Trial Steering Committee |
| WMH | White Matter Hyperintensity |

#### REFERENCES

1. Wardlaw J, Bath PMW, Doubal F, et al. Protocol: The Lacunar Intervention Trial 2 (LACI-2). A trial of two repurposed licenced drugs to prevent progression of cerebral small vessel disease. *Eur Stroke J* 2020;5(3):297-308. doi: 10.1177/2396987320920110 [published Online First: 20200420]
2. Gamble C, Krishan A, Stocken D, et al. Guidelines for the Content of Statistical Analysis Plans in Clinical Trials. *JAMA* 2017;318(23):2337-43. doi: 10.1001/jama.2017.18556
3. Hemming K, Kearney A, Gamble C, et al. Prospective reporting of statistical analysis plans for randomised controlled trials. *Trials* 2020;21(1):898. doi: 10.1186/s13063-020-04828-8 [published Online First: 20201028]
4. MacMahon S, Collins R. Reliable assessment of the effects of treatment on mortality and major morbidity, II: observational studies. *Lancet* 2001;357(9254):455-62. doi: 10.1016/s0140-6736(00)04017-4 [published Online First: 2001/03/29]
5. SPS3 Investigators BO, Hart RG, McClure LA, Szychowski JM, Coffey CS, Pearce LA. Effects of clopidogrel added to asprin in patients with recent lacunar stroke. *New England Journal of Medicine* 2012;376(9):817-25. doi: 10.1056/NEJMoa1204133
6. Bath PM, Woodhouse L, Scutt P, et al. Efficacy of nitric oxide, with or without continuing antihypertensive treatment, for management of high blood pressure in acute stroke (ENOS): a partial-factorial randomised controlled trial. *Lancet* 2015;385(9968):617-28. doi: 10.1016/s0140-6736(14)61121-1 [published Online First: 2014/12/04]
7. Appleton JP, Woodhouse LJ, Adami A, et al. Imaging markers of small vessel disease and brain frailty, and outcomes in acute stroke. *Neurology* 2020;94(5):e439-e52. doi: 10.1212/WNL.0000000000008881 [published Online First: 2019/12/27]
8. Sandercock P, Wardlaw JM, Lindley RI, et al. The benefits and harms of intravenous thrombolysis with recombinant tissue plasminogen activator within 6 h of acute ischaemic stroke (the third international stroke trial [IST-3]): a randomised controlled trial. *Lancet* 2012;379(9834):2352-63. doi: 10.1016/S0140-6736(12)60768-5 [published Online First: 20120523]
9. Sandercock P, Wardlaw JM, Dennis M, et al. Effect of thrombolysis with alteplase within 6 h of acute ischaemic stroke on long-term outcomes (the third International Stroke Trial IST-3 ): 18-month follow-up of a randomised controlled trial. *Lancet Neurology* 2013;12(8):768-76. doi: 10.1016/s1474-4422(13)70130-3
10. Schmidt R, Berghold A, Jokinen H, et al. White matter lesion progression in LADIS: frequency, clinical effects, and sample size calculations. *Stroke* 2012;43(10):2643-7. doi: 10.1161/strokeaha.112.662593 [published Online First: 2012/08/11]
11. McHutchison CA, Cvoro V, Makin S, et al. Functional, cognitive and physical outcomes 3 years after minor lacunar or cortical ischaemic stroke. *J Neurol Neurosurg Psychiatry* 2019;90(4):436-43. doi: 10.1136/jnnp-2018-319134 [published Online First: 20181215]
12. Wardlaw JM, Doubal FN, Valdes-Hernandez M, et al. Blood-brain barrier permeability and long-term clinical and imaging outcomes in cerebral small vessel disease. *Stroke* 2013;44(2):525-7. doi: 10.1161/strokeaha.112.669994 [published Online First: 2012/12/13]
13. Chappell FM, Del Carmen Valdés Hernández M, Makin SD, et al. Sample size considerations for trials using cerebral white matter hyperintensity progression as an intermediate outcome at 1 year after mild stroke: results of a prospective cohort study. *Trials* 2017;18(1):78. doi: 10.1186/s13063-017-1825-7 [published Online First: 20170221]
14. Pavlovic AM, Pekmezovic T, Tomic G, et al. Baseline predictors of cognitive decline in patients with cerebral small vessel disease. *Journal of Alzheimer's disease : JAD* 2014;42 Suppl 3:S37-43. doi: 10.3233/JAD-132606

15. Staals J, Makin SD, Doubal FN, et al. Stroke subtype, vascular risk factors, and total MRI brain small-vessel disease burden. *Neurology* 2014;83(14):1228-34. doi: 10.1212/WNL.0000000000000837 [published Online First: 20140827]
16. Lees RA, Hendry Ba K, Broomfield N, et al. Cognitive assessment in stroke: feasibility and test properties using differing approaches to scoring of incomplete items. *Int J Geriatr Psychiatry* 2017;32(10):1072-78. doi: 10.1002/gps.4568 [published Online First: 2016/08/16]
17. Bath PM, Woodhouse LJ, Appleton JP, et al. Antiplatelet therapy with aspirin, clopidogrel, and dipyridamole versus clopidogrel alone or aspirin and dipyridamole in patients with acute cerebral ischaemia (TARDIS): a randomised, open-label, phase 3 superiority trial. *Lancet* 2018;391(10123):850-59. doi: 10.1016/s0140-6736(17)32849-0 [published Online First: 2017/12/25]
18. Lachin J. Applications of the Wei-Lachin multivariate one-sided test for multiple outcomes on possibly different scales. *PLoS One* 2014;9(10) doi: 10.1371/journal.pone.0108784
19. Muresanu D, Heiss W, Hoemberg V, et al. Cerebrolysin and Recovery After Stroke (CARS): A Randomized, Placebo-Controlled, Double-Blind, Multicenter Trial. *Stroke* 2016;47(1):151-9. doi: 10.1161/STROKEAHA.115.009416 [published Online First: 12 Nov 2015]
20. Appleton J, Scutt P, Sprigg N, et al. Hypercholesterolaemia and vascular dementia. *Clinical Sciences* 2017;131(14):1561-78. doi: 10.1042/CS20160382 [published Online First: 21 March 2017]
21. Bath PM, Woodhouse LJ, Krishnan K, et al. Prehospital Transdermal Glyceryl Trinitrate for Ultra-Acute Intracerebral Hemorrhage: Data From the RIGHT-2 Trial. *Stroke* 2019;50(11):3064-71. doi: 10.1161/STROKEAHA.119.026389 [published Online First: 2019/10/07]
22. Sachdev PS, Blacker D, Blazer DG, et al. Classifying neurocognitive disorders: the DSM-5 approach. *Nature reviews Neurology* 2014;10(11):634-42. doi: 10.1038/nrneurol.2014.181 [published Online First: 2014/09/30]
23. Wardlaw JM, Doubal F, Brown R, et al. Rates, risks and routes to reduce vascular dementia (R4vad), a UK-wide multicentre prospective observational cohort study of cognition after stroke: Protocol. *European Stroke Journal* 2021;6(1):89-101. doi: 10.1177/2396987320953312
24. Wardlaw J, Bath P, Sandercock P, et al. The NeuroGrid stroke examplar clinical trial protocol. *International Journal of Stroke* 2007;2:63-9.

#### Main paper tables

##### Table 1. Baseline characteristics by treatment group isosorbide mononitrate (ISMN), cilostazol (Cil) or both (ISMN+Cil).

Data are number (%), median [interquartile range] or mean (standard deviation).

| | N | All | ISMN | No ISMN | Cil | No Cil | ISMN + Cil | ISMN only | Cil only | Neither |
|---|---|---|---|---|---|---|---|---|---|---|
| N | XX | XX | XX | XX | XX | XX | XX | XX | XX | XX |
| **Demographics** | | | | | | | | | | |
| Age (yr) † | XX | XX (X) | XX (X) | XX (X) | XX (X) | XX (X) | XX (X) | XX (X) | XX (X) | XX (X) |
| <=70 years | XX | XX (X) | XX (X) | XX (X) | XX | XX (X) | XX | XX (X) | XX (X) | XX (X) |
| Sex, female (%) | XX | XX (X) | XX (X) | XX (X) | XX (X) | XX (X) | XX (X) | XX (X) | XX (X) | XX (X) |
| modified Rankin Scale >1 (%) † | XX | XX (X) | XX (X) | XX (X) | XX (X) | XX (X) | XX (X) | XX (X) | XX (X) | XX (X) |
| Onset to randomisation (days) † | XX | XX (X) | XX (X) | XX (X) | XX (X) | XX (X) | XX (X) | XX (X) | XX (X) | XX (X) |
| <= 100 days | XX | XX (X) | XX (X) | XX (X) | XX (X) | XX (X) | XX (X) | XX (X) | XX (X) | XX (X) |
| Age completing education (yr) | XX | XX (X) | XX (X) | XX (X) | XX (X) | XX (X) | XX (X) | XX (X) | XX (X) | XX (X) |
| Highest education (%) † | | | | | | | | | | |
| Primary | XX | XX (X) | XX (X) | XX (X) | XX (X) | XX (X) | XX (X) | XX (X) | XX (X) | XX (X) |
| Secondary | XX | XX (X) | XX (X) | XX (X) | XX (X) | XX (X) | XX (X) | XX (X) | XX (X) | XX (X) |
| O' level/GCSE | XX | XX (X) | XX (X) | XX (X) | XX (X) | XX (X) | XX (X) | XX (X) | XX (X) | XX (X) |
| A' level | XX | XX (X) | XX (X) | XX (X) | XX (X) | XX (X) | XX (X) | XX (X) | XX (X) | XX (X) |
| Undergraduate degree | XX | XX (X) | XX (X) | XX (X) | XX (X) | XX (X) | XX (X) | XX (X) | XX (X) | XX (X) |
| Postgraduate degree | XX | XX (X) | XX (X) | XX (X) | XX (X) | XX (X) | XX (X) | XX (X) | XX (X) | XX (X) |
| ***Lifestyle*** | | | | | | | | | | |
| Smoking † | | | | | | | | | | |
| Current | XX | XX (X) | XX (X) | XX (X) | XX (X) | XX (X) | XX (X) | XX (X) | XX (X) | XX (X) |
| Past | XX | XX (X) | XX (X) | XX (X) | XX (X) | XX (X) | XX (X) | XX (X) | XX (X) | XX (X) |
| Never | XX | XX (X) | XX (X) | XX (X) | XX (X) | XX (X) | XX (X) | XX (X) | XX (X) | XX (X) |
| **History (%)** | | | | | | | | | | |
| Hypertension, drug treated | XX | XX (X) | XX (X) | XX (X) | XX (X) | XX (X) | XX (X) | XX (X) | XX (X) | XX (X) |
| Hyperlipidaemia, drug treated | XX | XX (X) | XX (X) | XX (X) | XX (X) | XX (X) | XX (X) | XX (X) | XX (X) | XX (X) |
| Diabetes mellitus | | | | | | | | | | |
| Oral agents | XX | XX (X) | XX (X) | XX (X) | XX (X) | XX (X) | XX (X) | XX (X) | XX (X) | XX (X) |
| Insulin | XX | XX (X) | XX (X) | XX (X) | XX (X) | XX (X) | XX (X) | XX (X) | XX (X) | XX (X) |
| Atrial fibrillation | XX | XX (X) | XX (X) | XX (X) | XX (X) | XX (X) | XX (X) | XX (X) | XX (X) | XX (X) |
| Heart failure | XX | XX (X) | XX (X) | XX (X) | XX (X) | XX (X) | XX (X) | XX (X) | XX (X) | XX (X) |
| Previous stroke | XX | XX (X) | XX (X) | XX (X) | XX (X) | XX (X) | XX (X) | XX (X) | XX (X) | XX (X) |
| Previous TIA | XX | XX (X) | XX (X) | XX (X) | XX (X) | XX (X) | XX (X) | XX (X) | XX (X) | XX (X) |

| | | | | | | | | | | |
|---|---|---|---|---|---|---|---|---|---|---|
| Family history, young stroke | XX | XX (X) | XX (X) | XX (X) | XX (X) | XX (X) | XX (X) | XX (X) | XX (X) | XX (X) |
| **Medications** | | | | | | | | | | |
| Anticoagulants | XX | XX (X) | XX (X) | XX (X) | XX (X) | XX (X) | XX (X) | XX (X) | XX (X) | XX (X) |
| Antibiotics | XX | XX (X) | XX (X) | XX (X) | XX (X) | XX (X) | XX (X) | XX (X) | XX (X) | XX (X) |
| Antihypertensives | XX | XX (X) | XX (X) | XX (X) | *XX (X)* | *XX (X)* | *XX (X)* | *XX (X)* | *XX (X)* | *XX (X)* |
| Antiplatelets | XX | XX (X) | XX (X) | XX (X) | *XX (X)* | *XX (X)* | *XX (X)* | XX (X) | XX (X) | XX (X) |
| Lipid-lowering | XX | XX (X) | XX (X) | XX (X) | *XX (X)* | *XX (X)* | *XX (X)* | XX (X) | XX (X) | XX (X) |
| Proton pump inhibitor | XX | XX (X) | XX (X) | XX (X) | *XX (X)* | *XX (X)* | XX (X) | XX (X) | XX (X) | XX (X) |
| PDE5 inhibitor | XX | XX (X) | XX (X) | XX (X) | *XX (X)* | *XX (X)* | XX (X) | XX (X) | XX (X) | XX (X) |
| Other drugs | XX | XX (X) | XX (X) | XX (X) | *XX (X)* | *XX (X)* | XX (X) | XX (X) | XX (X) | XX (X) |
| No. medications /day | XX | XX (X) | XX (X) | XX (X) | XX (X) | XX (X) | XX (X) | XX (X) | XX (X) | XX (X) |
| Grapefruit juice (%) | XX | XX (X) | XX (X) | XX (X) | XX (X) | XX (X) | XX (X) | XX (X) | XX (X) | XX (X) |
| **Clinical (%)** | | | | | | | | | | |
| Systolic BP (mmHg) † | XX | XX (X) | XX (X) | XX (X) | XX (X) | XX (X) | XX (X) | XX (X) | XX (X) | XX (X) |
| Diastolic BP mmHg) | XX | XX (X) | XX (X) | XX (X) | XX (X) | XX (X) | XX (X) | XX (X) | XX (X) | XX (X) |
| Atrial fibrillation | XX | XX (X) | XX (X) | XX (X) | *XX (X)* | *XX (X)* | *XX (X)* | *XX (X)* | *XX (X)* | *XX (X)* |
| NIHSS (/42) † | XX | XX (X) | XX (X) | XX (X) | XX (X) | XX (X) | XX (X) | XX (X) | XX (X) | XX (X) |
| Weakness, Side (%) | | | | | | | | | | |
| right | XX | XX (X) | XX (X) | XX (X) | XX (X) | XX (X) | XX (X) | XX (X) | XX (X) | XX (X) |
| left | XX | XX (X) | XX (X) | XX (X) | XX (X) | XX (X) | XX (X) | XX (X) | XX (X) | XX (X) |
| both | XX | XX (X) | XX (X) | XX (X) | XX (X) | XX (X) | XX (X) | XX (X) | XX (X) | XX (X) |
| Sensory loss (%) | | | | | | | | | | |
| Right | XX | XX (X) | XX (X) | XX (X) | XX (X) | XX (X) | XX (X) | XX (X) | XX (X) | XX (X) |
| Left | XX | XX (X) | XX (X) | XX (X) | XX (X) | XX (X) | XX (X) | XX (X) | XX (X) | XX (X) |
| both | XX | XX (X) | XX (X) | XX (X) | XX (X) | XX (X) | XX (X) | XX (X) | XX (X) | XX (X) |
| Ataxia (%) | | | | | | | | | | |
| left | XX | XX (X) | XX (X) | XX (X) | XX | XX (X) | XX (X) | XX (X) | XX | XX (X) |
| right | XX | XX (X) | XX (X) | XX (X) | XX | XX (X) | XX (X) | XX (X) | XX | XX (X) |
| Neglect/inattention (%) | XX | XX (X) | XX (X) | XX (X) | XX (X) | XX (X) | XX (X) | XX (X) | XX (X) | XX (X) |
| Dysphasia (%) | XX | XX (X) | XX (X) | XX (X) | XX (X) | XX (X) | XX (X) | XX (X) | XX (X) | XX (X) |
| Dysarthria (%) | XX | XX (X) | XX (X) | XX (X) | XX (X) | XX (X) | XX (X) | XX (X) | XX (X) | XX (X) |
| Visual loss (%) | XX | XX (X) | XX (X) | XX (X) | XX (X) | XX (X) | XX (X) | XX (X) | XX (X) | XX (X) |
| **Cognition** | | | | | | | | | | |
| MoCA | XX | XX (X) | XX (X) | XX (X) | XX (X) | XX (X) | XX (X) | XX (X) | XX (X) | XX (X) |
| MoCA <=24 | XX | XX (X) | XX (X) | XX (X) | | | XX (X) | XX (X) | XX (X) | |
| Verbal fluency F, <11 words | XX | XX (X) | XX (X) | XX (X) | XX (X) | XX (X) | XX (X) | XX (X) | XX (X) | XX (X) |
| Trails B, time | XX | XX (X) | XX (X) | XX (X) | XX (X) | XX (X) | XX (X) | XX (X) | XX (X) | XX (X) |
| Trails B, points | XX | XX (X) | XX (X) | XX (X) | XX (X) | XX (X) | XX (X) | XX (X) | XX (X) | XX (X) |
| **Investigations** | | | | | | | | | | |
| CT scan | XX | XX (X) | XX (X) | XX (X) | XX (X) | XX (X) | XX (X) | XX (X) | XX (X) | XX (X) |
| MRI Scan | XX | XX (X) | XX (X) | XX (X) | XX (X) | XX (X) | XX (X) | XX (X) | XX (X) | XX (X) |
| Both scans | XX | XX (X) | XX (X) | XX (X) | XX (X) | XX (X) | XX (X) | XX (X) | XX (X) | XX (X) |
| Stroke-CT scan (days) | XX | XX (X) | XX (X) | XX (X) | XX (X) | XX (X) | XX (X) | XX (X) | XX (X) | XX (X) |
| Stroke-MRI scan (days) | XX | XX (X) | XX (X) | XX (X) | XX (X) | XX (X) | XX (X) | XX (X) | XX (X) | XX (X) |

| | | | | | | | | | | |
|---|---|---|---|---|---|---|---|---|---|---|
| Index infarct present (%) | XX | XX (X) | XX (X) | XX (X) | XX (X) | XX (X) | XX (X) | XX (X) | XX (X) | XX (X) |
| Index infarct side, left (%) | XX | XX (X) | XX (X) | XX (X) | XX (X) | XX (X) | XX (X) | XX (X) | XX (X) | XX (X) |
| cSVD moderate/severe (%) | XX | XX (X) | XX (X) | XX (X) | XX (X) | XX (X) | XX (X) | XX (X) | XX (X) | XX (X) |
| WMH/ hypoattenuations | XX | XX (X) | XX (X) | XX (X) | XX (X) | XX (X) | XX (X) | XX (X) | XX (X) | XX (X) |
| Carotid stenosis | XX | XX (X) | XX (X) | XX (X) | | | XX (X) | XX (X) | | |
| Left >=50% | XX | XX (X) | XX (X) | XX (X) | XX (X) | XX (X) | XX (X) | XX (X) | XX (X) | XX (X) |
| Right >=50% | XX | XX (X) | XX (X) | XX (X) | XX (X) | XX (X) | XX (X) | XX (X) | XX (X) | XX (X) |
| ECG, (%) | | | | | | | | | | |
| Sinus | XX | XX (X) | XX (X) | XX (X) | XX (X) | XX (X) | XX (X) | XX (X) | XX (X) | XX (X) |
| AF | XX | XX (X) | XX (X) | XX (X) | XX (X) | XX (X) | XX (X) | XX (X) | XX (X) | XX (X) |
| Haemoglobin (g/l) | XX | XX (X) | XX (X) | XX (X) | XX (X) | XX (X) | XX (X) | XX (X) | XX (X) | XX (X) |
| Creatinine (μmol/l) | XX | XX (X) | XX (X) | XX (X) | XX (X) | XX (X) | XX (X) | XX (X) | XX (X) | XX (X) |
| eGFR (ml/min) | XX | XX (X) | XX (X) | XX (X) | XX (X) | XX (X) | XX (X) | XX (X) | XX (X) | XX (X) |
| **Contraindications to treatment** | | | | | | | | | | |
| ISMN | XX | XX (X) | XX (X) | XX (X) | XX (X) | XX (X) | XX (X) | XX (X) | XX (X) | XX (X) |
| Cilostazol | XX | XX (X) | XX (X) | XX (X) | XX (X) | XX (X) | XX (X) | XX (X) | XX (X) | XX (X) |

† Minimisation variable

ECG: electrocardiogram; F/T: full time; ICH: intracerebral haemorrhage; IS: ischaemic stroke; LACS: lacunar syndrome; LVH: left ventricular hypertrophy; PACS; partial anterior circulation syndrome; POCS: posterior circulation syndrome; P/T: part time; TACS: total anterior circulation syndrome; TIA: transient ischaemic attack

**Table 2. Feasibility measures.**
Data are number (%).

| Measure | Metric | Achieved |
|---|---|---|
| ***Primary*** | | |
| Recruitment | 400 patients | 363/400 (90.8%) |
| Retention of enrolees at 1 year | >95% | XX (X) |
| ***Secondary*** | | |
| Tolerability | >=75% on at least half dose | XX (X) |
| ISMN alone | | XX (X) |
| Cilostazol alone | | XX (X) |
| Both ISMN and cilostazol | | XX (X) |
| **Safety** | | |
| Symptomatic extracranial bleeding | | XX (X) |
| Symptomatic intracranial bleeding | | XX (X) |
| Death | <2% | XX (X) |
| Stroke | | XX (X) |
| Haemorrhage | | XX (X) |
| Extracranial | | XX (X) |
| Intracranial | | XX (X) |
| **Efficacy** | | XX (X) |
| Stroke | | XX (X) |
| TIA | | XX (X) |
| Myocardial infarction | | XX (X) |
| Cognitive impairment | | XX (X) |
| Dependency, mRS>2 | | XX (X) |
| Any of these | 40-50% | XX (X) |

**Table 3. Clinical Outcomes at 12 months.**
Data are number (%), median [interquartile range] or mean (standard deviation). Analyses performed using binary logistic regression (BLR), ordinal logistic regression (OLR) or multiple linear regression (MLR) with adjustment for age, sex, time from stroke onset to randomisation, years of education, smoking status, and baseline mRS (dependency), stroke severity (NIHSS) and systolic blood pressure. Mean (SD) and MLR will be used instead of median [IQR] and OLR for ordinal scales with more than 7 levels (central limit theorem/large sample). The Wei-Lachin test is used to analyse multiple outcomes in parallel.

| Number | **ISMN** | **No ISMN** | **Difference (p)** | **Cilostazol** | **No cilostazol** | **Difference (p)** | **ISMN + Cil** | **Neither** | **Difference (p)** |
|---|---|---|---|---|---|---|---|---|---|
| **Composite** | | | CPHR | | | CPHR | | | CPHR |
| Stroke | XX (X) | XX (X) | BLR | XX (X) | XX (X) | BLR | XX (X) | XX (X) | BLR |
| TIA | XX (X) | XX (X) | BLR | XX (X) | XX (X) | BLR | XX (X) | XX (X) | BLR |
| MI | XX (X) | XX (X) | BLR | XX (X) | XX (X) | BLR | XX (X) | XX (X) | BLR |
| Cognitive impairment | XX (X) | XX (X) | BLR | XX (X) | XX (X) | BLR | XX (X) | XX (X) | BLR |
| Dependency, mRS>2 | XX (X) | XX (X) | BLR | XX (X) | XX (X) | BLR | XX (X) | XX (X) | BLR |
| Death | XX (X) | XX (X) | BLR | XX (X) | XX (X) | BLR | XX (X) | XX (X) | BLR |
| **Cognition** | | | | | | | | | |
| Cognition, 7 level | | | OLR | | | OLR | | | OLR |
| Normal | XX (X) | XX (X) | | XX (X) | XX (X) | | XX (X) | XX (X) | |
| Minor, single domain | XX (X) | XX (X) | | XX (X) | XX (X) | | XX (X) | XX (X) | |
| Minor, multi-domain | XX (X) | XX (X) | | XX (X) | XX (X) | | XX (X) | XX (X) | |
| Major, mild | XX (X) | XX (X) | | XX (X) | XX (X) | | XX (X) | XX (X) | |

| | | | | | | | | | |
|---|---|---|---|---|---|---|---|---|---|
| Major, moderate | XX (X) | XX (X) | | XX (X) | XX (X) | | XX (X) | XX (X) | |
| Major, severe | XX (X) | XX (X) | | XX (X) | XX (X) | | XX (X) | XX (X) | |
| Death | XX (X) | XX (X) | | XX (X) | XX (X) | | XX (X) | XX (X) | |
| Cognition, 4 level | | | OLR | | | OLR | | | OLR |
| Normal | XX (X) | XX (X) | | XX (X) | XX (X) | | XX (X) | XX (X) | |
| Minor | XX (X) | XX (X) | | XX (X) | XX (X) | | XX (X) | XX (X) | |
| Dementia | XX (X) | XX (X) | | XX (X) | XX (X) | | XX (X) | XX (X) | |
| Death | XX (X) | XX (X) | | XX (X) | XX (X) | | XX (X) | XX (X) | |
| Memory/thinking problem | XX (X) | XX (X) | BLR | XX (X) | XX (X) | BLR | XX (X) | XX (X) | BLR |
| MoCA | XX (X) | XX (X) | MLR | XX (X) | XX (X) | MLR | XX (X) | XX (X) | MLR |
| TICS-m | XX (X) | XX (X) | MLR | XX (X) | XX (X) | MLR | XX (X) | XX (X) | MLR |
| Verbal fluency, animal naming | XX (X) | XX (X) | MLR | XX (X) | XX (X) | MLR | XX (X) | XX (X) | MLR |
| Trails B, time | XX (X) | XX (X) | MLR | XX (X) | XX (X) | MLR | XX (X) | XX (X) | MLR |
| Trails B, points | XX (X) | XX (X) | MLR | XX (X) | XX (X) | MLR | XX (X) | XX (X) | MLR |
| Dementia, clinical diagnosis | XX (X) | XX (X) | BLR | XX (X) | XX (X) | BLR | XX (X) | XX (X) | BLR |
| **Clinical** | | | | | | | | | |
| Systolic BP (mmHg) | XX (X) | XX (X) | MLR | XX (X) | XX (X) | MLR | XX (X) | XX (X) | MLR |
| Diastolic BP (mmHg) | XX (X) | XX (X) | MLR | XX (X) | XX (X) | MLR | XX (X) | XX (X) | MLR |

| | | | | | | | | | |
|---|---|---|---|---|---|---|---|---|---|
| Heart rate (bpm) | XX (X) | XX (X) | MLR | XX (X) | XX (X) | MLR | XX (X) | XX (X) | MLR |
| mRS | XX (X) | XX (X) | OLR | XX (X) | XX (X) | OLR | XX (X) | XX (X) | OLR |
| Disposition | XX (X) | XX (X) | OLR | XX (X) | XX (X) | OLR | XX (X) | XX (X) | OLR |
| ZDS | XX (X) | XX (X) | MLR | XX (X) | XX (X) | MLR | XX (X) | XX (X) | MLR |
| Clinical depression | XX (X) | XX (X) | BLR | XX (X) | XX (X) | BLR | XX (X) | XX (X) | BLR |
| EQ-5D-5L, as HU | XX (X) | XX (X) | MLR | XX (X) | XX (X) | MLR | XX (X) | XX (X) | MLR |
| EQ-VAS | XX (X) | XX (X) | MLR | XX (X) | XX (X) | MLR | XX (X) | XX (X) | MLR |
| SIS | XX (X) | XX (X) | MLR | XX (X) | XX (X) | MLR | XX (X) | XX (X) | MLR |
| Global | XX (X) | XX (X) | WLT | XX (X) | XX (X) | WLT | XX (X) | XX (X) | WLT |

BP: blood pressure; MoCA: Montreal cognitive assessment-modified; mRS: modified Rankin Scale; SIS: stroke impact scale; TICS-m: Telephone interview cognitive status- modified; ZDS: Zung depression scale
Global: Recurrent ordinal stroke,[17] ordinal MI, cognition (MoCA), dependency (mRS), stroke impact scale, quality of life (EQ-5D).

**Table 4. Adherence to medication with at least half dose or more by randomised group: isosorbide mononitrate (ISMN), cilostazol (Cil) and both together.**

| Week | ISMN | No ISMN | p | Cilostazol | No cilostazol | p | ISMN +Cil | ISMN alone | Cil only | Neither | p |
|---|---|---|---|---|---|---|---|---|---|---|---|
| 1-2 | XX (X) | XX (X) | Chi-sq | XX (X) | XX (X) | Chi-sq | XX (X) | XX (X) | XX (X) | XX (X) | Chi-sq |
| 3-4 | XX (X) | XX (X) | Chi-sq | XX (X) | XX (X) | Chi-sq | XX (X) | XX (X) | XX (X) | XX (X) | Chi-sq |
| 26 | XX (X) | XX (X) | Chi-sq | XX (X) | XX (X) | Chi-sq | XX (X) | XX (X) | XX (X) | XX (X) | Chi-sq |
| 52 | XX (X) | XX (X) | Chi-sq | XX (X) | XX (X) | Chi-sq | XX (X) | XX (X) | XX (X) | XX (X) | Chi-sq |

A more detailed table by strata of drug adherence (i.e. 25%, 50%, 75% and 100% adherence) will also be prepared.

**Table 5. Adjudicated baseline imaging characteristics by randomised group: isosorbide mononitrate (ISMN), cilostazol (Cil) and both together.**
Data are number (%), median [IQR], or mean (standard deviation).

| | N | All | ISMN | No ISMN | Cil | No Cil | ISMN + Cil | ISMN only | Cil only | None |
|---|---|---|---|---|---|---|---|---|---|---|
| Patients randomised | XX | XX | XX | XX | XX | XX | XX | XX | XX | XX |
| **Scan** | | | | | | | | | | |
| *Scan type(%)* | | | | | | | | | | |
| CT | XX | XX (X) | XX (X) | XX (X) | XX (X) | XX (X) | XX (X) | XX (X) | XX (X) | XX (X) |
| Time to scan (days) | XX | XX (X) | XX (X) | XX (X) | XX (X) | XX (X) | XX (X) | XX (X) | XX (X) | XX (X) |
| MR | XX | XX (X) | XX (X) | XX (X) | XX (X) | XX (X) | XX (X) | XX (X) | XX (X) | XX (X) |
| Time to scan (days) | XX | XX (X) | XX (X) | XX (X) | XX (X) | XX (X) | XX (X) | XX (X) | XX (X) | XX (X) |
| Scan quality (%) | | | | | | | | | | |
| Good | XX | XX (X) | XX (X) | XX (X) | XX (X) | XX (X) | XX (X) | XX (X) | XX (X) | XX (X) |
| Moderate | XX | XX (X) | XX (X) | XX (X) | XX (X) | XX (X) | XX (X) | XX (X) | XX (X) | XX (X) |
| *Poor* | XX | XX (X) | XX (X) | XX (X) | XX (X) | XX (X) | XX (X) | XX (X) | XX (X) | XX (X) |
| **Index Lesion (i.e. main cause of stroke symptoms) (%)** | | | | | | | | | | |
| Normal Scan | XX | XX (X) | XX (X) | XX (X) | XX (X) | XX (X) | XX (X) | XX (X) | XX (X) | XX (X) |
| Lesion present (type) | | | | | | | | | | |
| Primary Acute ischaemia | XX | XX (X) | XX (X) | XX (X) | XX (X) | XX (X) | XX (X) | XX (X) | XX (X) | XX (X) |
| Primary haemorrhage | XX | XX (X) | XX (X) | XX (X) | XX (X) | XX (X) | XX (X) | XX (X) | XX (X) | XX (X) |
| Mimic | XX | XX (X) | XX (X) | XX (X) | XX (X) | XX (X) | XX (X) | XX (X) | XX (X) | XX (X) |
| No visible | XX | XX (X) | XX (X) | XX (X) | XX (X) | XX (X) | XX (X) | XX (X) | XX (X) | XX (X) |
| Infarct side of brain | | | | | | | | | | |
| Right | XX | XX (X) | XX (X) | XX (X) | XX (X) | XX (X) | XX (X) | XX (X) | XX (X) | XX (X) |
| Left | XX | XX (X) | XX (X) | XX (X) | XX (X) | XX (X) | XX (X) | XX (X) | XX (X) | XX (X) |
| Both | XX | XX (X) | XX (X) | XX (X) | XX (X) | XX (X) | XX (X) | XX (X) | XX (X) | XX (X) |
| **Location (%)** | | | | | | | | | | |

| | N | All | ISMN | No ISMN | Cil | No Cil | ISMN + Cil | ISMN only | Cil only | None |
|---|---|---|---|---|---|---|---|---|---|---|
| *Index small subcortical (i.e. acute lacunar) infarct* | XX | XX (X) | XX (X) | XX (X) | XX (X) | XX (X) | XX (X) | XX (X) | XX (X) | XX (X) |
| Internal capsule | XX | XX (X) | XX (X) | XX (X) | XX (X) | XX (X) | XX (X) | XX (X) | XX (X) | XX (X) |
| External capsule | XX | XX (X) | XX (X) | XX (X) | XX (X) | XX (X) | XX (X) | XX (X) | XX (X) | XX (X) |
| Lentiform nucleus | XX | XX (X) | XX (X) | XX (X) | XX (X) | XX (X) | XX (X) | XX (X) | XX (X) | XX (X) |
| Internal border zone | XX | XX (X) | XX (X) | XX (X) | XX (X) | XX (X) | XX (X) | XX (X) | XX (X) | XX (X) |
| Centrum semiovale | XX | XX (X) | XX (X) | XX (X) | XX (X) | XX (X) | XX (X) | XX (X) | XX (X) | XX (X) |
| Thalamus | XX | XX (X) | XX (X) | XX (X) | XX (X) | XX (X) | XX (X) | XX (X) | XX (X) | XX (X) |
| Lacunar- small deep cerebellar lesion | XX | XX (X) | XX (X) | XX (X) | XX (X) | XX (X) | XX (X) | XX (X) | XX (X) | XX (X) |
| Lacunar- small deep brainstem lesion (Pons) | XX | XX (X) | XX (X) | XX (X) | XX (X) | XX (X) | XX (X) | XX (X) | XX (X) | XX (X) |
| Lacunar - Medulla | XX | XX (X) | XX (X) | XX (X) | XX (X) | XX (X) | XX (X) | XX (X) | XX (X) | XX (X) |
| **Non-small subcortical (i.e. large artery cortical or large subcortical or posterior circulation) infarct** | | | | | | | | | | |
| MCA territory | XX | XX (X) | XX (X) | XX (X) | XX (X) | XX (X) | XX (X) | XX (X) | XX (X) | XX (X) |
| PCA territory | XX | XX (X) | XX (X) | XX (X) | XX (X) | XX (X) | XX (X) | XX (X) | XX (X) | XX (X) |
| **Infarct size (mm)** | | | | | | | | | | |
| A/P | XX | XX (X) | XX (X) | XX (X) | XX (X) | XX (X) | XX (X) | XX (X) | XX (X) | XX (X) |
| R/L | XX | XX (X) | XX (X) | XX (X) | XX (X) | XX (X) | XX (X) | XX (X) | XX (X) | XX (X) |
| Cranio-caudal | XX | XX (X) | XX (X) | XX (X) | XX (X) | XX (X) | XX (X) | XX (X) | XX (X) | XX (X) |
| **Microhaemorrhage (%)** | | | | | | | | | | |
| Microhaemorrhages | XX | XX (X) | XX (X) | XX (X) | XX (X) | XX (X) | XX (X) | XX (X) | XX (X) | XX (X) |
| *No. microhaemorrhages (%)* | | | | | | | | | | |
| 1 | XX | XX (X) | XX (X) | XX (X) | XX (X) | XX (X) | XX (X) | XX (X) | XX (X) | XX (X) |
| 2 | XX | XX (X) | XX (X) | XX (X) | XX (X) | XX (X) | XX (X) | XX (X) | XX (X) | XX (X) |
| 3 | XX | XX (X) | XX (X) | XX (X) | XX (X) | XX (X) | XX (X) | XX (X) | XX (X) | XX (X) |
| 4 | XX | XX (X) | XX (X) | XX (X) | XX (X) | XX (X) | XX (X) | XX (X) | XX (X) | XX (X) |
| >= 5 | XX | XX (X) | XX (X) | XX (X) | XX (X) | XX (X) | XX (X) | XX (X) | XX (X) | XX (X) |
| *Type of microhaemorrhages (%)* | | | | | | | | | | |

| | N | All | ISMN | No ISMN | Cil | No Cil | ISMN + Cil | ISMN only | Cil only | None |
|---|---|---|---|---|---|---|---|---|---|---|
| Lobar | XX | XX (X) | XX (X) | XX (X) | XX (X) | XX (X) | XX (X) | XX (X) | XX (X) | XX (X) |
| Deep | XX | XX (X) | XX (X) | XX (X) | XX (X) | XX (X) | XX (X) | XX (X) | XX (X) | XX (X) |
| Both | XX | XX (X) | XX (X) | XX (X) | XX (X) | XX (X) | XX (X) | XX (X) | XX (X) | XX (X) |
| Superficial siderosis present | XX | XX (X) | XX (X) | XX (X) | XX (X) | XX (X) | XX (X) | XX (X) | XX (X) | XX (X) |
| Siderosis focal | XX | XX (X) | XX (X) | XX (X) | XX (X) | XX (X) | XX (X) | XX (X) | XX (X) | XX (X) |
| Siderosis disseminated | XX | XX (X) | XX (X) | XX (X) | XX (X) | XX (X) | XX (X) | XX (X) | XX (X) | XX (X) |
| *Siderosis location* | | | | | | | | | | |
| Left | XX | XX (X) | XX (X) | XX (X) | XX (X) | XX (X) | XX (X) | XX (X) | XX (X) | XX (X) |
| Right | XX | XX (X) | XX (X) | XX (X) | XX (X) | XX (X) | XX (X) | XX (X) | XX (X) | XX (X) |
| Both | XX | XX (X) | XX (X) | XX (X) | XX (X) | XX (X) | XX (X) | XX (X) | XX (X) | XX (X) |
| **Atrophy** | | | | | | | | | | |
| Brain tissue volume reduction | XX | XX (X) | XX (X) | XX (X) | XX (X) | XX (X) | XX (X) | XX (X) | XX (X) | XX (X) |
| Central brain tissue volume | | | | | | | | | | |
| Modest | XX | XX (X) | XX (X) | XX (X) | XX (X) | XX (X) | XX (X) | XX (X) | XX (X) | XX (X) |
| Severe | XX | XX (X) | XX (X) | XX (X) | XX (X) | XX (X) | XX (X) | XX (X) | XX (X) | XX (X) |
| Cortical brain tissue volume | | | | | | | | | | |
| Modest | XX | XX (X) | XX (X) | XX (X) | XX (X) | XX (X) | XX (X) | XX (X) | XX (X) | XX (X) |
| Severe | XX | XX (X) | XX (X) | XX (X) | XX (X) | XX (X) | XX (X) | XX (X) | XX (X) | XX (X) |
| **White Matter hyperintensities (%)** | | | | | | | | | | |
| White matter hyperintensities | XX | XX (X) | XX (X) | XX (X) | XX (X) | XX (X) | XX (X) | XX (X) | XX (X) | XX (X) |
| *Anterior white matter lucency* | | | | | | | | | | |
| Restricted region adjoining ventricles | XX | XX (X) | XX (X) | XX (X) | XX (X) | XX (X) | XX (X) | XX (X) | XX (X) | XX (X) |
| Covering ventricle to cortex | XX | XX (X) | XX (X) | XX (X) | XX (X) | XX (X) | XX (X) | XX (X) | XX (X) | XX (X) |
| *Posterior white matter lucency* | | | | | | | | | | |
| Restricted region adjoining ventricles | XX | XX (X) | XX (X) | XX (X) | XX (X) | XX (X) | XX (X) | XX (X) | XX (X) | XX (X) |
| Covering ventricle to cortex | XX | XX (X) | XX (X) | XX (X) | XX (X) | XX (X) | XX (X) | XX (X) | XX (X) | XX (X) |
| *Anterior and/or Posterior white matter lucency* | | | | | | | | | | |

| | N | All | ISMN | No ISMN | Cil | No Cil | ISMN + Cil | ISMN only | Cil only | None |
|---|---|---|---|---|---|---|---|---|---|---|
| Restricted region adjoining ventricles | XX | XX (X) | XX (X) | XX (X) | XX (X) | XX (X) | XX (X) | XX (X) | XX (X) | XX (X) |
| Covering ventricle to cortex | XX | XX (X) | XX (X) | XX (X) | XX (X) | XX (X) | XX (X) | XX (X) | XX (X) | XX (X) |
| *Periventricular WMH Fazekas score* | | | | | | | | | | |
| 1 | XX | XX (X) | XX (X) | XX (X) | XX (X) | XX (X) | XX (X) | XX (X) | XX (X) | XX (X) |
| 2 | XX | XX (X) | XX (X) | XX (X) | XX (X) | XX (X) | XX (X) | XX (X) | XX (X) | XX (X) |
| 3 | XX | XX (X) | XX (X) | XX (X) | XX (X) | XX (X) | XX (X) | XX (X) | XX (X) | XX (X) |
| *Deep WMH Fazekas score* | | | | | | | | | | |
| 1 | XX | XX (X) | XX (X) | XX (X) | XX (X) | XX (X) | XX (X) | XX (X) | XX (X) | XX (X) |
| 2 | XX | XX (X) | XX (X) | XX (X) | XX (X) | XX (X) | XX (X) | XX (X) | XX (X) | XX (X) |
| 3 | XX | XX (X) | XX (X) | XX (X) | XX (X) | XX (X) | XX (X) | XX (X) | XX (X) | XX (X) |
| *Periventricular and/or Deep WMH Fazekas score* | | | | | | | | | | |
| 1 | XX | XX (X) | XX (X) | XX (X) | XX (X) | XX (X) | XX (X) | XX (X) | XX (X) | XX (X) |
| 2 | XX | XX (X) | XX (X) | XX (X) | XX (X) | XX (X) | XX (X) | XX (X) | XX (X) | XX (X) |
| 3 | XX | XX (X) | XX (X) | XX (X) | XX (X) | XX (X) | XX (X) | XX (X) | XX (X) | XX (X) |
| **Enlarged perivascular spaces (%)** | | | | | | | | | | |
| Enlarged perivascular spaces | XX | XX (X) | XX (X) | XX (X) | XX (X) | XX (X) | XX (X) | XX (X) | XX (X) | XX (X) |
| *Basal ganglia rating, worse side* | | | | | | | | | | |
| <=10 | XX | XX (X) | XX (X) | XX (X) | XX (X) | XX (X) | XX (X) | XX (X) | XX (X) | XX (X) |
| 11-20 | XX | XX (X) | XX (X) | XX (X) | XX (X) | XX (X) | XX (X) | XX (X) | XX (X) | XX (X) |
| 20-40 | XX | XX (X) | XX (X) | XX (X) | XX (X) | XX (X) | XX (X) | XX (X) | XX (X) | XX (X) |
| >40 | XX | XX (X) | XX (X) | XX (X) | XX (X) | XX (X) | XX (X) | XX (X) | XX (X) | XX (X) |
| *Centrum semiovale rating, worse side* | | | | | | | | | | |
| <=10 | XX | XX (X) | XX (X) | XX (X) | XX (X) | XX (X) | XX (X) | XX (X) | XX (X) | XX (X) |
| 11-20 | XX | XX (X) | XX (X) | XX (X) | XX (X) | XX (X) | XX (X) | XX (X) | XX (X) | XX (X) |
| 20-40 | XX | XX (X) | XX (X) | XX (X) | XX (X) | XX (X) | XX (X) | XX (X) | XX (X) | XX (X) |
| >40 | XX | XX (X) | XX (X) | XX (X) | XX (X) | XX (X) | XX (X) | XX (X) | XX (X) | XX (X) |
| **Old vascular lesions (%)** | | | | | | | | | | |

| | N | All | ISMN | No ISMN | Cil | No Cil | ISMN + Cil | ISMN only | Cil only | None |
|---|---|---|---|---|---|---|---|---|---|---|
| Old vascular lesions | XX | XX (X) | XX (X) | XX (X) | XX (X) | XX (X) | XX (X) | XX (X) | XX (X) | XX (X) |
| Old cortical infarct | XX | XX (X) | XX (X) | XX (X) | XX (X) | XX (X) | XX (X) | XX (X) | XX (X) | XX (X) |
| Old striatocapsular infarct | XX | XX (X) | XX (X) | XX (X) | XX (X) | XX (X) | XX (X) | XX (X) | XX (X) | XX (X) |
| Old borderzone infarct | XX | XX (X) | XX (X) | XX (X) | XX (X) | XX (X) | XX (X) | XX (X) | XX (X) | XX (X) |
| Old lacunar infarct | XX | XX (X) | XX (X) | XX (X) | XX (X) | XX (X) | XX (X) | XX (X) | XX (X) | XX (X) |
| *Number of lacunes (%)* | | | | | | | | | | |
| 1 | XX | XX (X) | XX (X) | XX (X) | XX (X) | XX (X) | XX (X) | XX (X) | XX (X) | XX (X) |
| 2 | XX | XX (X) | XX (X) | XX (X) | XX (X) | XX (X) | XX (X) | XX (X) | XX (X) | XX (X) |
| 3 | XX | XX (X) | XX (X) | XX (X) | XX (X) | XX (X) | XX (X) | XX (X) | XX (X) | XX (X) |
| 4 | XX | XX (X) | XX (X) | XX (X) | XX (X) | XX (X) | XX (X) | XX (X) | XX (X) | XX (X) |
| >=5 | XX | XX (X) | XX (X) | XX (X) | XX (X) | XX (X) | XX (X) | XX (X) | XX (X) | XX (X) |
| Old brainstem/cerebellar infarcts | XX | XX (X) | XX (X) | XX (X) | XX (X) | XX (X) | XX (X) | XX (X) | XX (X) | XX (X) |
| Probable old haemorrhage | XX | XX (X) | XX (X) | XX (X) | XX (X) | XX (X) | XX (X) | XX (X) | XX (X) | XX (X) |
| **Non-stroke lesions (%)** | | | | | | | | | | |
| Non-stroke lesion | XX | XX (X) | XX (X) | XX (X) | XX (X) | XX (X) | XX (X) | XX (X) | XX (X) | XX (X) |
| *Classification of non-stroke (%)* | | | | | | | | | | |
| Cerebral tumour | XX | XX (X) | XX (X) | XX (X) | XX (X) | XX (X) | XX (X) | XX (X) | XX (X) | XX (X) |
| Aneurysm | XX | XX (X) | XX (X) | XX (X) | XX (X) | XX (X) | XX (X) | XX (X) | XX (X) | XX (X) |
| Vascular malformation | XX | XX (X) | XX (X) | XX (X) | XX (X) | XX (X) | XX (X) | XX (X) | XX (X) | XX (X) |
| Other non-stroke classification | XX | XX (X) | XX (X) | XX (X) | XX (X) | XX (X) | XX (X) | XX (X) | XX (X) | XX (X) |

**Table 6a. Serious adverse events for ISMN.**
Data are number (%).

| | All | | | Fatal | | |
|---|---|---|---|---|---|---|
| Number | ISMN | No ISMN | Difference (p) | ISMN | No ISMN | Difference (p) |
| **Treatment** | | | | | | |
| During | xx (xx.x) | xx (xx.x) | BLR | xx (xx.x) | xx (xx.x) | BLR |
| After | xx (xx.x) | xx (xx.x) | BLR | | | BLR |
| **Relationship** | | | | | | |
| Possibly | xx (xx.x) | xx (xx.x) | Ch sq | xx (xx.x) | xx (xx.x) | Ch sq |
| Probably | xx (xx.x) | xx (xx.x) | - | xx (xx.x) | xx (xx.x) | - |
| Definitely | xx (xx.x) | xx (xx.x) | - | xx (xx.x) | xx (xx.x) | - |

**Table 6b. Serious adverse events for cilostazol.**
Data are number (%).

| | All | | | Fatal | | |
|---|---|---|---|---|---|---|
| Number | Cilostazol | No Cilostazol | Difference (p) | Cilostazol | No Cilostazol | Difference (p) |
| **Treatment** | | | | | | |
| During | xx (xx.x) | xx (xx.x) | BLR | xx (xx.x) | xx (xx.x) | BLR |
| After | xx (xx.x) | xx (xx.x) | BLR | xx (xx.x) | xx (xx.x) | BLR |
| **Relationship** | | | | | | |
| Possibly | xx (xx.x) | xx (xx.x) | Ch sq | xx (xx.x) | xx (xx.x) | Ch sq |
| Probably | xx (xx.x) | xx (xx.x) | - | xx (xx.x) | xx (xx.x) | - |
| Definitely | xx (xx.x) | xx (xx.x) | - | xx (xx.x) | xx (xx.x) | - |

**Table 6c. Serious adverse events for combined ISMN and cilostazol.**
Data are number (%).

| | All | | | Fatal | | |
|---|---|---|---|---|---|---|
| Number | ISMN /cil | No ISMN/cil | Difference (p) | ISMN /cil | No ISMN/cil | Difference (p) |
| **Treatment** | | | | | | |
| During | xx (xx.x) | xx (xx.x) | BLR | xx (xx.x) | xx (xx.x) | BLR |
| After | xx (xx.x) | xx (xx.x) | BLR | xx (xx.x) | xx (xx.x) | BLR |
| **Relationship** | | | | | | |
| Possibly | xx (xx.x) | xx (xx.x) | Ch sq | xx (xx.x) | xx (xx.x) | Ch sq |
| Probably | xx (xx.x) | xx (xx.x) | - | xx (xx.x) | xx (xx.x) | - |
| Definitely | xx (xx.x) | xx (xx.x) | - | xx (xx.x) | xx (xx.x) | - |

**Table 7a. Participants with at least one serious adverse events by organ randomised to isosorbide mononitrate (ISMN) versus none.**
Data are number (%); comparison by binary logistic regression.

| Number | All ISMN | All No ISMN | All Difference (p) | Fatal ISMN | Fatal No ISMN | Fatal Difference (p) |
|---|---|---|---|---|---|---|
| Cardiovascular | xx (xx.x) | xx (xx.x) | (xx.x, xx.x), p=0.xxx | xx (xx.x) | xx (xx.x) | (xx.x, xx.x), p=0.xxx |
| Myocardial infarction | xx (xx.x) | xx (xx.x) | (xx.x, xx.x), p=0.xxx | xx (xx.x) | xx (xx.x) | (xx.x, xx.x), p=0.xxx |
| Nervous system | xx (xx.x) | xx (xx.x) | (xx.x, xx.x), p=0.xxx | xx (xx.x) | xx (xx.x) | (xx.x, xx.x), p=0.xxx |
| Ischaemic stroke | xx (xx.x) | xx (xx.x) | (xx.x, xx.x), p=0.xxx | xx (xx.x) | xx (xx.x) | (xx.x, xx.x), p=0.xxx |
| Transient ischaemic attack | xx (xx.x) | xx (xx.x) | (xx.x, xx.x), p=0.xxx | xx (xx.x) | xx (xx.x) | (xx.x, xx.x), p=0.xxx |
| Intracerebral haemorrhage | xx (xx.x) | xx (xx.x) | (xx.x, xx.x), p=0.xxx | xx (xx.x) | xx (xx.x) | (xx.x, xx.x), p=0.xxx |
| Respiratory | xx (xx.x) | xx (xx.x) | (xx.x, xx.x), p=0.xxx | xx (xx.x) | xx (xx.x) | (xx.x, xx.x), p=0.xxx |
| Gastrointestinal | xx (xx.x) | xx (xx.x) | (xx.x, xx.x), p=0.xxx | xx (xx.x) | xx (xx.x) | (xx.x, xx.x), p=0.xxx |
| Genitourinary | xx (xx.x) | xx (xx.x) | (xx.x, xx.x), p=0.xxx | xx (xx.x) | xx (xx.x) | (xx.x, xx.x), p=0.xxx |
| Secondary | xx (xx.x) | xx (xx.x) | (xx.x, xx.x), p=0.xxx | xx (xx.x) | xx (xx.x) | (xx.x, xx.x), p=0.xxx |
| Haematological | xx (xx.x) | xx (xx.x) | (xx.x, xx.x), p=0.xxx | xx (xx.x) | xx (xx.x) | (xx.x, xx.x), p=0.xxx |
| Metabolic/endocrine | xx (xx.x) | xx (xx.x) | (xx.x, xx.x), p=0.xxx | xx (xx.x) | xx (xx.x) | (xx.x, xx.x), p=0.xxx |
| Musculoskeletal | xx (xx.x) | xx (xx.x) | (xx.x, xx.x), p=0.xxx | xx (xx.x) | xx (xx.x) | (xx.x, xx.x), p=0.xxx |
| Infection/sepsis | xx (xx.x) | xx (xx.x) | (xx.x, xx.x), p=0.xxx | xx (xx.x) | xx (xx.x) | (xx.x, xx.x), p=0.xxx |
| Tumour/malignancy | xx (xx.x) | xx (xx.x) | (xx.x, xx.x), p=0.xxx | xx (xx.x) | xx (xx.x) | (xx.x, xx.x), p=0.xxx |
| Other | xx (xx.x) | xx (xx.x) | (xx.x, xx.x), p=0.xxx | xx (xx.x) | xx (xx.x) | (xx.x, xx.x), p=0.xxx |
| Total | xx (xx.x) | xx (xx.x) | (xx.x, xx.x), p=0.xxx | xx (xx.x) | xx (xx.x) | (xx.x, xx.x), p=0.xxx |

**Table 7b. Participants with at least one serious adverse events by organ randomised to cilostazol (Cil) versus none.**
Data are number (%); comparison by binary logistic regression.

| Number | All<br>Cil | <br>No Cil | <br>Difference (p) | Fatal<br>Cil | <br>No Cil | <br>Difference (p) |
|---|---|---|---|---|---|---|
| Cardiovascular | xx (xx.x) | xx (xx.x) | (xx.x, xx.x), p=0.xxx | xx (xx.x) | xx (xx.x) | (xx.x, xx.x), p=0.xxx |
| Myocardial infarction | xx (xx.x) | xx (xx.x) | (xx.x, xx.x), p=0.xxx | xx (xx.x) | xx (xx.x) | (xx.x, xx.x), p=0.xxx |
| Nervous system | xx (xx.x) | xx (xx.x) | (xx.x, xx.x), p=0.xxx | xx (xx.x) | xx (xx.x) | (xx.x, xx.x), p=0.xxx |
| Ischaemic stroke | xx (xx.x) | xx (xx.x) | (xx.x, xx.x), p=0.xxx | xx (xx.x) | xx (xx.x) | (xx.x, xx.x), p=0.xxx |
| Transient ischaemic attack | xx (xx.x) | xx (xx.x) | (xx.x, xx.x), p=0.xxx | xx (xx.x) | xx (xx.x) | (xx.x, xx.x), p=0.xxx |
| Intracerebral haemorrhage | xx (xx.x) | xx (xx.x) | (xx.x, xx.x), p=0.xxx | xx (xx.x) | xx (xx.x) | (xx.x, xx.x), p=0.xxx |
| Respiratory | xx (xx.x) | xx (xx.x) | (xx.x, xx.x), p=0.xxx | xx (xx.x) | xx (xx.x) | (xx.x, xx.x), p=0.xxx |
| Gastrointestinal | xx (xx.x) | xx (xx.x) | (xx.x, xx.x), p=0.xxx | xx (xx.x) | xx (xx.x) | (xx.x, xx.x), p=0.xxx |
| Genitourinary | xx (xx.x) | xx (xx.x) | (xx.x, xx.x), p=0.xxx | xx (xx.x) | xx (xx.x) | (xx.x, xx.x), p=0.xxx |
| Secondary | xx (xx.x) | xx (xx.x) | (xx.x, xx.x), p=0.xxx | xx (xx.x) | xx (xx.x) | (xx.x, xx.x), p=0.xxx |
| Haematological | xx (xx.x) | xx (xx.x) | (xx.x, xx.x), p=0.xxx | xx (xx.x) | xx (xx.x) | (xx.x, xx.x), p=0.xxx |
| Metabolic/endocrine | xx (xx.x) | xx (xx.x) | (xx.x, xx.x), p=0.xxx | xx (xx.x) | xx (xx.x) | (xx.x, xx.x), p=0.xxx |
| Musculoskeletal | xx (xx.x) | xx (xx.x) | (xx.x, xx.x), p=0.xxx | xx (xx.x) | xx (xx.x) | (xx.x, xx.x), p=0.xxx |
| Infection/sepsis | xx (xx.x) | xx (xx.x) | (xx.x, xx.x), p=0.xxx | xx (xx.x) | xx (xx.x) | (xx.x, xx.x), p=0.xxx |
| Tumour/malignancy | xx (xx.x) | xx (xx.x) | (xx.x, xx.x), p=0.xxx | xx (xx.x) | xx (xx.x) | (xx.x, xx.x), p=0.xxx |
| Other | xx (xx.x) | xx (xx.x) | (xx.x, xx.x), p=0.xxx | xx (xx.x) | xx (xx.x) | (xx.x, xx.x), p=0.xxx |
| Total | xx (xx.x) | xx (xx.x) | (xx.x, xx.x), p=0.xxx | xx (xx.x) | xx (xx.x) | (xx.x, xx.x), p=0.xxx |

**Table 7c. Participants with at least one serious adverse events by organ randomised to combined isosorbide mononitrate (ISMN) and cilostazol (Cil) versus neither.**

Data are number (%); comparison by binary logistic regression.

| | All | | | Fatal | | |
|---|---|---|---|---|---|---|
| Number | **ISMN /Cil** | **No ISNM /Cil** | **Difference (p)** | **ISMN /Cil** | **No ISNM /Cil** | **Difference (p)** |
| Cardiovascular | xx (xx.x) | xx (xx.x) | (xx.x, xx.x), p=0.xxx | xx (xx.x) | xx (xx.x) | (xx.x, xx.x), p=0.xxx |
| Myocardial infarction | xx (xx.x) | xx (xx.x) | (xx.x, xx.x), p=0.xxx | xx (xx.x) | xx (xx.x) | (xx.x, xx.x), p=0.xxx |
| Nervous system | xx (xx.x) | xx (xx.x) | (xx.x, xx.x), p=0.xxx | xx (xx.x) | xx (xx.x) | (xx.x, xx.x), p=0.xxx |
| Ischaemic stroke | xx (xx.x) | xx (xx.x) | (xx.x, xx.x), p=0.xxx | xx (xx.x) | xx (xx.x) | (xx.x, xx.x), p=0.xxx |
| Transient ischaemic attack | xx (xx.x) | xx (xx.x) | (xx.x, xx.x), p=0.xxx | xx (xx.x) | xx (xx.x) | (xx.x, xx.x), p=0.xxx |
| Intracerebral haemorrhage | xx (xx.x) | xx (xx.x) | (xx.x, xx.x), p=0.xxx | xx (xx.x) | xx (xx.x) | (xx.x, xx.x), p=0.xxx |
| Respiratory | xx (xx.x) | xx (xx.x) | (xx.x, xx.x), p=0.xxx | xx (xx.x) | xx (xx.x) | (xx.x, xx.x), p=0.xxx |
| Gastrointestinal | xx (xx.x) | xx (xx.x) | (xx.x, xx.x), p=0.xxx | xx (xx.x) | xx (xx.x) | (xx.x, xx.x), p=0.xxx |
| Genitourinary | xx (xx.x) | xx (xx.x) | (xx.x, xx.x), p=0.xxx | xx (xx.x) | xx (xx.x) | (xx.x, xx.x), p=0.xxx |
| Secondary | xx (xx.x) | xx (xx.x) | (xx.x, xx.x), p=0.xxx | xx (xx.x) | xx (xx.x) | (xx.x, xx.x), p=0.xxx |
| Haematological | xx (xx.x) | xx (xx.x) | (xx.x, xx.x), p=0.xxx | xx (xx.x) | xx (xx.x) | (xx.x, xx.x), p=0.xxx |
| Metabolic/endocrine | xx (xx.x) | xx (xx.x) | (xx.x, xx.x), p=0.xxx | xx (xx.x) | xx (xx.x) | (xx.x, xx.x), p=0.xxx |
| Musculoskeletal | xx (xx.x) | xx (xx.x) | (xx.x, xx.x), p=0.xxx | xx (xx.x) | xx (xx.x) | (xx.x, xx.x), p=0.xxx |
| Infection/sepsis | xx (xx.x) | xx (xx.x) | (xx.x, xx.x), p=0.xxx | xx (xx.x) | xx (xx.x) | (xx.x, xx.x), p=0.xxx |
| Tumour/malignancy | xx (xx.x) | xx (xx.x) | (xx.x, xx.x), p=0.xxx | xx (xx.x) | xx (xx.x) | (xx.x, xx.x), p=0.xxx |
| Other | xx (xx.x) | xx (xx.x) | (xx.x, xx.x), p=0.xxx | xx (xx.x) | xx (xx.x) | (xx.x, xx.x), p=0.xxx |
| Total | xx (xx.x) | xx (xx.x) | (xx.x, xx.x), p=0.xxx | xx (xx.x) | xx (xx.x) | (xx.x, xx.x), p=0.xxx |

**Table 8. Targeted symptoms occurring at any time on treatment on isosorbide mononitrate (ISMN), cilostazol (cil) or both.**
Data are number (%); comparison by binary logistic regression.

| | ISMN | No ISMN | Difference (p) | Cil | No cil | Difference (p) | ISMN /Cil | No ISMN /Cil | Difference (p) |
|---|---|---|---|---|---|---|---|---|---|
| Headache | xx (xx.x) | xx (xx.x) | (xx.x, xx.x), p=0.xxx | xx (xx.x) | xx (xx.x) | (xx.x, xx.x), p=0.xxx | xx (xx.x) | xx (xx.x) | (xx.x, xx.x), p=0.xxx |
| Stopped normal activities | xx (xx.x) | xx (xx.x) | (xx.x, xx.x), p=0.xxx | xx (xx.x) | xx (xx.x) | (xx.x, xx.x), p=0.xxx | xx (xx.x) | xx (xx.x) | (xx.x, xx.x), p=0.xxx |
| Palpitations | xx (xx.x) | xx (xx.x) | (xx.x, xx.x), p=0.xxx | xx (xx.x) | xx (xx.x) | (xx.x, xx.x), p=0.xxx | xx (xx.x) | xx (xx.x) | (xx.x, xx.x), p=0.xxx |
| Stopped normal activities | xx (xx.x) | xx (xx.x) | (xx.x, xx.x), p=0.xxx | xx (xx.x) | xx (xx.x) | (xx.x, xx.x), p=0.xxx | xx (xx.x) | xx (xx.x) | (xx.x, xx.x), p=0.xxx |
| Dizziness | xx (xx.x) | xx (xx.x) | (xx.x, xx.x), p=0.xxx | xx (xx.x) | xx (xx.x) | (xx.x, xx.x), p=0.xxx | xx (xx.x) | xx (xx.x) | (xx.x, xx.x), p=0.xxx |
| Stopped normal activities | xx (xx.x) | xx (xx.x) | (xx.x, xx.x), p=0.xxx | xx (xx.x) | xx (xx.x) | (xx.x, xx.x), p=0.xxx | xx (xx.x) | xx (xx.x) | (xx.x, xx.x), p=0.xxx |
| Loose stools | xx (xx.x) | xx (xx.x) | (xx.x, xx.x), p=0.xxx | xx (xx.x) | xx (xx.x) | (xx.x, xx.x), p=0.xxx | xx (xx.x) | xx (xx.x) | (xx.x, xx.x), p=0.xxx |
| Stopped normal activities | xx (xx.x) | xx (xx.x) | (xx.x, xx.x), p=0.xxx | xx (xx.x) | xx (xx.x) | (xx.x, xx.x), p=0.xxx | xx (xx.x) | xx (xx.x) | (xx.x, xx.x), p=0.xxx |
| Nausea | xx (xx.x) | xx (xx.x) | (xx.x, xx.x), p=0.xxx | xx (xx.x) | xx (xx.x) | (xx.x, xx.x), p=0.xxx | xx (xx.x) | xx (xx.x) | (xx.x, xx.x), p=0.xxx |
| Stopped normal activities | xx (xx.x) | xx (xx.x) | (xx.x, xx.x), p=0.xxx | xx (xx.x) | xx (xx.x) | (xx.x, xx.x), p=0.xxx | xx (xx.x) | xx (xx.x) | (xx.x, xx.x), p=0.xxx |
| Bleeding | xx (xx.x) | xx (xx.x) | (xx.x, xx.x), p=0.xxx | xx (xx.x) | xx (xx.x) | (xx.x, xx.x), p=0.xxx | xx (xx.x) | xx (xx.x) | (xx.x, xx.x), p=0.xxx |
| Stopped normal activities | xx (xx.x) | xx (xx.x) | (xx.x, xx.x), p=0.xxx | xx (xx.x) | xx (xx.x) | (xx.x, xx.x), p=0.xxx | xx (xx.x) | xx (xx.x) | (xx.x, xx.x), p=0.xxx |
| Bruising | xx (xx.x) | xx (xx.x) | (xx.x, xx.x), p=0.xxx | xx (xx.x) | xx (xx.x) | (xx.x, xx.x), p=0.xxx | xx (xx.x) | xx (xx.x) | (xx.x, xx.x), p=0.xxx |
| Stopped normal activities | xx (xx.x) | xx (xx.x) | (xx.x, xx.x), p=0.xxx | xx (xx.x) | xx (xx.x) | (xx.x, xx.x), p=0.xxx | xx (xx.x) | xx (xx.x) | (xx.x, xx.x), p=0.xxx |
| Falls | xx (xx.x) | xx (xx.x) | (xx.x, xx.x), p=0.xxx | xx (xx.x) | xx (xx.x) | (xx.x, xx.x), p=0.xxx | xx (xx.x) | xx (xx.x) | (xx.x, xx.x), p=0.xxx |
| Stopped normal activities | xx (xx.x) | xx (xx.x) | (xx.x, xx.x), p=0.xxx | xx (xx.x) | xx (xx.x) | (xx.x, xx.x), p=0.xxx | xx (xx.x) | xx (xx.x) | (xx.x, xx.x), p=0.xxx |

**Table 9. Adjudicated 1 year MRI imaging characteristics by randomised group: isosorbide mononitrate (ISMN), cilostazol (Cil) and both together.**
Data are number (%), median [IQR], or mean (standard deviation).

| | N | All | ISMN | No ISMN | OR/MD/ HR (95%CI) | p-value | Cil | No Cil | OR/MD/ HR (95%CI) | p-value | ISMN + Cil | None | OR/MD/ HR (95%CI) | p-value |
|---|---|---|---|---|---|---|---|---|---|---|---|---|---|---|
| Patients randomised | XX | XX | XX | XX | | | XX | XX | | | XX | XX | | |
| **Scan** | | | | | | | | | | | | | | |
| Time to scan (days) | XX | XX (X) | XX (X) | XX (X) | MLR | XX | XX (X) | XX (X) | MLR | XX | XX (X) | XX (X) | MLR | XX |
| Scan quality (%) | | | | | OLR | XX | | | OLR | XX | | | OLR | XX |
| Good | XX | XX (X) | XX (X) | XX (X) | | | XX (X) | XX (X) | | | XX (X) | XX (X) | | |
| Moderate | XX | XX (X) | XX (X) | XX (X) | | | XX (X) | XX (X) | | | XX (X) | XX (X) | | |
| *Poor* | XX | XX (X) | XX (X) | XX (X) | | | XX (X) | XX (X) | | | XX (X) | XX (X) | | |
| **Appearance of the index infarct now (%)** | | | | | | | | | | | | | | |
| Completely cavitated - visible on T2, FLAIR and T1" | XX | XX (X) | XX (X) | XX (X) | BLR | XX | XX (X) | XX (X) | BLR | XX | XX (X) | XX (X) | BLR | XX |
| Partially cavitated - lacy | XX | XX (X) | XX (X) | XX (X) | BLR | XX | XX (X) | XX (X) | BLR | XX | XX (X) | XX (X) | BLR | XX |
| Partially cavitated - hole + large WMH rim | XX | XX (X) | XX (X) | XX (X) | BLR | XX | XX (X) | XX (X) | BLR | XX | XX (X) | XX (X) | BLR | XX |
| Partially cavitated - FLAIR cavity but not=CSF | XX | XX (X) | XX (X) | XX (X) | BLR | XX | XX (X) | XX (X) | BLR | XX | XX (X) | XX (X) | BLR | XX |
| Partially cavitated - FLAIR=WMH T2=cavity | XX | XX (X) | XX (X) | XX (X) | BLR | XX | XX (X) | XX (X) | BLR | XX | XX (X) | XX (X) | BLR | XX |
| Partially cavitated - visible on T2 not FLAIR | XX | XX (X) | XX (X) | XX (X) | BLR | XX | XX (X) | XX (X) | BLR | XX | XX (X) | XX (X) | BLR | XX |
| Not cavitated (WML-like) | XX | XX (X) | XX (X) | XX (X) | BLR | XX | XX (X) | XX (X) | BLR | XX | XX (X) | XX (X) | BLR | XX |
| Disappeared | XX | XX (X) | XX (X) | XX (X) | BLR | XX | XX (X) | XX (X) | BLR | XX | XX (X) | XX (X) | BLR | XX |
| Become visible | XX | XX (X) | XX (X) | XX (X) | BLR | XX | XX (X) | XX (X) | BLR | XX | XX (X) | XX (X) | BLR | XX |
| Never visible | XX | XX (X) | XX (X) | XX (X) | BLR | XX | XX (X) | XX (X) | BLR | XX | XX (X) | XX (X) | BLR | XX |
| **Evidence of new stroke (%)** | | | | | | | | | | | | | | |

| | N | All | ISMN | No ISMN | OR/MD/ HR (95%CI) | p-value | Cil | No Cil | OR/MD/ HR (95%CI) | p-value | ISMN + Cil | None | OR/MD/ HR (95%CI) | p-value |
|---|---|---|---|---|---|---|---|---|---|---|---|---|---|---|
| Ischaemic | XX | XX (X) | XX (X) | XX (X) | BLR | XX | XX (X) | XX (X) | BLR | XX | XX (X) | XX (X) | BLR | XX |
| Haemorrhagic | XX | XX (X) | XX (X) | XX (X) | BLR | XX | XX (X) | XX (X) | BLR | XX | XX (X) | XX (X) | BLR | XX |
| **Microhaemorrhages (%)** | XX | XX (X) | XX (X) | XX (X) | BLR | XX | XX (X) | XX (X) | BLR | XX | XX (X) | XX (X) | BLR | XX |
| *No. microhaemorrhages (%)* | | | | | OLR | XX | | | OLR | XX | | | OLR | XX |
| 1 | XX | XX (X) | XX (X) | XX (X) | | | XX (X) | XX (X) | | | XX (X) | XX (X) | | |
| 2 | XX | XX (X) | XX (X) | XX (X) | | | XX (X) | XX (X) | | | XX (X) | XX (X) | | |
| 3 | XX | XX (X) | XX (X) | XX (X) | | | XX (X) | XX (X) | | | XX (X) | XX (X) | | |
| 4 | XX | XX (X) | XX (X) | XX (X) | | | XX (X) | XX (X) | | | XX (X) | XX (X) | | |
| >= 5 | XX | XX (X) | XX (X) | XX (X) | | | XX (X) | XX (X) | | | XX (X) | XX (X) | | |
| Change from baseline | XX | XX [XX, XX] | XX [XX, XX] | XX [XX, XX] | OLR | XX | XX [XX, XX] | XX [XX, XX] | OLR | XX | XX [XX, XX] | XX [XX, XX] | OLR | XX |
| **Small vessel disease score** | | | | | | | | | | | | | | |
| Total | XX | XX [XX, XX] | XX [XX, XX] | XX [XX, XX] | OLR | XX | XX [XX, XX] | XX [XX, XX] | OLR | XX | XX [XX, XX] | XX [XX, XX] | OLR | XX |
| Change from baseline | XX | XX [XX, XX] | XX [XX, XX] | XX [XX, XX] | OLR | XX | XX [XX, XX] | XX [XX, XX] | OLR | XX | XX [XX, XX] | XX [XX, XX] | OLR | XX |
| **Atrophy** | | | | | | | | | | | | | | |
| Brain tissue volume reduction | XX | XX (X) | XX (X) | XX (X) | BLR | XX | XX (X) | XX (X) | BLR | XX | XX (X) | XX (X) | BLR | XX |
| Central brain tissue volume | | | | | OLR | XX | | | OLR | XX | | | OLR | XX |
| Modest | XX | XX (X) | XX (X) | XX (X) | | | XX (X) | XX (X) | | | XX (X) | XX (X) | | |
| Severe | XX | XX (X) | XX (X) | XX (X) | | | XX (X) | XX (X) | | | XX (X) | XX (X) | | |
| Cortical brain tissue volume | | | | | OLR | XX | | | OLR | XX | | | OLR | XX |
| Modest | XX | XX (X) | XX (X) | XX (X) | | | XX (X) | XX (X) | | | XX (X) | XX (X) | | |
| Severe | XX | XX (X) | XX (X) | XX (X) | | | XX (X) | XX (X) | | | XX (X) | XX (X) | | |
| Change from baseline | | | | | OLR | XX | | | OLR | XX | | | OLR | XX |
| More | XX | XX (X) | XX (X) | XX (X) | | | XX (X) | XX (X) | | | XX (X) | XX (X) | | |
| Less | XX | XX (X) | XX (X) | XX (X) | | | XX (X) | XX (X) | | | XX (X) | XX (X) | | |
| None | XX | XX (X) | XX (X) | XX (X) | | | XX (X) | XX (X) | | | XX (X) | XX (X) | | |

| | N | All | ISMN | No ISMN | OR/MD/ HR (95%CI) | p-value | Cil | No Cil | OR/MD/ HR (95%CI) | p-value | ISMN + Cil | None | OR/MD/ HR (95%CI) | p-value |
|---|---|---|---|---|---|---|---|---|---|---|---|---|---|---|
| **White Matter hyperintensities (%)** | | | | | | | | | | | | | | |
| White matter hyperintensities | XX | XX (X) | XX (X) | XX (X) | BLR | XX | XX (X) | XX (X) | BLR | XX | XX (X) | XX (X) | BLR | XX |
| *Anterior white matter lucency* | | | | | | | | | | | | | | |
| Restricted region adjoining ventricles | XX | XX (X) | XX (X) | XX (X) | BLR | XX | XX (X) | XX (X) | BLR | XX | XX (X) | XX (X) | BLR | XX |
| Covering ventricle to cortex | XX | XX (X) | XX (X) | XX (X) | BLR | XX | XX (X) | XX (X) | BLR | XX | XX (X) | XX (X) | BLR | XX |
| *Posterior white matter lucency* | | | | | | | | | | | | | | |
| Restricted region adjoining ventricles | XX | XX (X) | XX (X) | XX (X) | BLR | XX | XX (X) | XX (X) | BLR | XX | XX (X) | XX (X) | BLR | XX |
| Covering ventricle to cortex | XX | XX (X) | XX (X) | XX (X) | BLR | XX | XX (X) | XX (X) | BLR | XX | XX (X) | XX (X) | BLR | XX |
| *Anterior and/or Posterior white matter lucency* | | | | | | | | | | | | | | |
| Restricted region adjoining ventricles | XX | XX (X) | XX (X) | XX (X) | BLR | XX | XX (X) | XX (X) | BLR | XX | XX (X) | XX (X) | BLR | XX |
| Covering ventricle to cortex | XX | XX (X) | XX (X) | XX (X) | BLR | XX | XX (X) | XX (X) | BLR | XX | XX (X) | XX (X) | BLR | XX |
| *Periventricular WMH Fazekas score* | | | | | OLR | XX | | | OLR | XX | | | OLR | XX |
| 1 | XX | XX (X) | XX (X) | XX (X) | | | XX (X) | XX (X) | | | XX (X) | XX (X) | | |
| 2 | XX | XX (X) | XX (X) | XX (X) | | | XX (X) | XX (X) | | | XX (X) | XX (X) | | |
| 3 | XX | XX (X) | XX (X) | XX (X) | | | XX (X) | XX (X) | | | XX (X) | XX (X) | | |
| Change from baseline | XX | XX [XX, XX] | XX [XX, XX] | XX [XX, XX] | OLR | XX | XX [XX, XX] | XX [XX, XX] | OLR | XX | XX [XX, XX] | XX [XX, XX] | OLR | XX |
| *Deep WMH Fazekas score* | | | | | OLR | XX | | | OLR | XX | | | OLR | XX |
| 1 | XX | XX (X) | XX (X) | XX (X) | | | XX (X) | XX (X) | | | XX (X) | XX (X) | | |
| 2 | XX | XX (X) | XX (X) | XX (X) | | | XX (X) | XX (X) | | | XX (X) | XX (X) | | |
| 3 | XX | XX (X) | XX (X) | XX (X) | | | XX (X) | XX (X) | | | XX (X) | XX (X) | | |
| Change from baseline | XX | XX [XX, XX] | XX [XX, XX] | XX [XX, XX] | OLR | XX | XX [XX, XX] | XX [XX, XX] | OLR | XX | XX [XX, XX] | XX [XX, XX] | OLR | XX |
| *Periventricular and/or Deep WMH Fazekas score* | | | | | OLR | XX | | | OLR | XX | | | OLR | XX |
| 1 | XX | XX (X) | XX (X) | XX (X) | | | XX (X) | XX (X) | | | XX (X) | XX (X) | | |
| 2 | XX | XX (X) | XX (X) | XX (X) | | | XX (X) | XX (X) | | | XX (X) | XX (X) | | |

| | N | All | ISMN | No ISMN | OR/MD/ HR (95%CI) | p-value | Cil | No Cil | OR/MD/ HR (95%CI) | p-value | ISMN + Cil | None | OR/MD/ HR (95%CI) | p-value |
|---|---|---|---|---|---|---|---|---|---|---|---|---|---|---|
| 3 | XX | XX (X) | XX (X) | XX (X) | | | XX (X) | XX (X) | | | XX (X) | XX (X) | | |
| Change from baseline | XX | XX [XX, XX] | XX [XX, XX] | XX [XX, XX] | OLR | XX | XX [XX, XX] | XX [XX, XX] | OLR | XX | XX [XX, XX] | XX [XX, XX] | OLR | XX |
| WMH change from randomisation | XX | XX (X) | XX (X) | XX (X) | BLR | XX | XX (X) | XX (X) | BLR | XX | XX (X) | XX (X) | BLR | XX |
| *Frontal (%)* | | | | | OLR | XX | | | OLR | XX | | | OLR | XX |
| More | XX | XX (X) | XX (X) | XX (X) | | | XX (X) | XX (X) | | | XX (X) | XX (X) | | |
| Less | XX | XX (X) | XX (X) | XX (X) | | | XX (X) | XX (X) | | | XX (X) | XX (X) | | |
| No Change | XX | XX (X) | XX (X) | XX (X) | | | XX (X) | XX (X) | | | XX (X) | XX (X) | | |
| *Parietal (%)* | | | | | OLR | XX | | | OLR | XX | | | OLR | XX |
| More | XX | XX (X) | XX (X) | XX (X) | | | XX (X) | XX (X) | | | XX (X) | XX (X) | | |
| Less | XX | XX (X) | XX (X) | XX (X) | | | XX (X) | XX (X) | | | XX (X) | XX (X) | | |
| No Change | XX | XX (X) | XX (X) | XX (X) | | | XX (X) | XX (X) | | | XX (X) | XX (X) | | |
| *Occipital (%)* | | | | | OLR | XX | | | OLR | XX | | | OLR | XX |
| More | XX | XX (X) | XX (X) | XX (X) | | | XX (X) | XX (X) | | | XX (X) | XX (X) | | |
| Less | XX | XX (X) | XX (X) | XX (X) | | | XX (X) | XX (X) | | | XX (X) | XX (X) | | |
| No Change | XX | XX (X) | XX (X) | XX (X) | | | XX (X) | XX (X) | | | XX (X) | XX (X) | | |
| *Basal ganglia (%)* | | | | | OLR | XX | | | OLR | XX | | | OLR | XX |
| More | XX | XX (X) | XX (X) | XX (X) | | | XX (X) | XX (X) | | | XX (X) | XX (X) | | |
| Less | XX | XX (X) | XX (X) | XX (X) | | | XX (X) | XX (X) | | | XX (X) | XX (X) | | |
| No Change | XX | XX (X) | XX (X) | XX (X) | | | XX (X) | XX (X) | | | XX (X) | XX (X) | | |
| *Posterior fossa (%)* | | | | | OLR | XX | | | OLR | XX | | | OLR | XX |
| More | XX | XX (X) | XX (X) | XX (X) | | | XX (X) | XX (X) | | | XX (X) | XX (X) | | |
| Less | XX | XX (X) | XX (X) | XX (X) | | | XX (X) | XX (X) | | | XX (X) | XX (X) | | |
| No Change | XX | XX (X) | XX (X) | XX (X) | | | XX (X) | XX (X) | | | XX (X) | XX (X) | | |
| **Old vascular lesions (%)** | | | | | | | | | | | | | | |
| Old vascular lesions | XX | XX (X) | XX (X) | XX (X) | BLR | XX | XX (X) | XX (X) | BLR | XX | XX (X) | XX (X) | BLR | XX |
| Old cortical infarct | XX | XX (X) | XX (X) | XX (X) | BLR | XX | XX (X) | XX (X) | BLR | XX | XX (X) | XX (X) | BLR | XX |
| Old striatocapsular infarct | XX | XX (X) | XX (X) | XX (X) | BLR | XX | XX (X) | XX (X) | BLR | XX | XX (X) | XX (X) | BLR | XX |

| | N | All | ISMN | No ISMN | OR/MD/ HR (95%CI) | p-value | Cil | No Cil | OR/MD/ HR (95%CI) | p-value | ISMN + Cil | None | OR/MD/ HR (95%CI) | p-value |
|---|---|---|---|---|---|---|---|---|---|---|---|---|---|---|
| Old borderzone infarct | XX | XX (X) | XX (X) | XX (X) | BLR | XX | XX (X) | XX (X) | BLR | XX | XX (X) | XX (X) | BLR | XX |
| Old lacunar infarct | XX | XX (X) | XX (X) | XX (X) | BLR | XX | XX (X) | XX (X) | BLR | XX | XX (X) | XX (X) | BLR | XX |
| *Number of lacunes (%)* | | | | | OLR | XX | | | OLR | XX | | | OLR | XX |
| 1 | XX | XX (X) | XX (X) | XX (X) | | | XX (X) | XX (X) | | | XX (X) | XX (X) | | |
| 2 | XX | XX (X) | XX (X) | XX (X) | | | XX (X) | XX (X) | | | XX (X) | XX (X) | | |
| 3 | XX | XX (X) | XX (X) | XX (X) | | | XX (X) | XX (X) | | | XX (X) | XX (X) | | |
| 4 | XX | XX (X) | XX (X) | XX (X) | | | XX (X) | XX (X) | | | XX (X) | XX (X) | | |
| >=5 | XX | XX (X) | XX (X) | XX (X) | | | XX (X) | XX (X) | | | XX (X) | XX (X) | | |
| Change in number of lacunes from baseline | XX | XX [XX, XX] | XX [XX, XX] | XX [XX, XX] | OLR | XX | XX [XX, XX] | XX [XX, XX] | OLR | XX | XX [XX, XX] | XX [XX, XX] | OLR | XX |
| Old brainstem/cerebellar infarcts | XX | XX (X) | XX (X) | XX (X) | BLR | XX | XX (X) | XX (X) | BLR | XX | XX (X) | XX (X) | BLR | XX |
| Probable old haemorrhage | XX | XX (X) | XX (X) | XX (X) | BLR | XX | XX (X) | XX (X) | BLR | XX | XX (X) | XX (X) | BLR | XX |
| **Non-stroke lesions (%)** | | | | | | | | | | | | | | |
| Non-stroke lesion | XX | XX (X) | XX (X) | XX (X) | BLR | XX | XX (X) | XX (X) | BLR | XX | XX (X) | XX (X) | BLR | XX |
| *Classification of non-stroke (%)* | | | | | | | | | | | | | | |
| Cerebral tumour | XX | XX (X) | XX (X) | XX (X) | BLR | XX | XX (X) | XX (X) | BLR | XX | XX (X) | XX (X) | BLR | XX |
| Aneurysm | XX | XX (X) | XX (X) | XX (X) | BLR | XX | XX (X) | XX (X) | BLR | XX | XX (X) | XX (X) | BLR | XX |
| Vascular malformation | XX | XX (X) | XX (X) | XX (X) | BLR | XX | XX (X) | XX (X) | BLR | XX | XX (X) | XX (X) | BLR | XX |
| Other non-stroke classification | XX | XX (X) | XX (X) | XX (X) | BLR | XX | XX (X) | XX (X) | BLR | XX | XX (X) | XX (X) | BLR | XX |

**Figures**

1. CONSORT flowchart diagram
2. Forest plot of composite outcome (stroke, TIA, MI, cognitive impairment, dependency or death) by clinical and adjudicated imaging subgroups
3. Forest plot of 7-level ordinal cognition/dementia scale by clinical and adjudicated imaging subgroups
4. Forest plot of Wei-Lachin global outcome by clinical and adjudicated imaging subgroups
5. Forest plot of central imaging reads as per list for table 9 of absolute and change in imaging findings
6. Stacked distributions of 7-level ordinal cognition at 12 months
7. Stacked distributions of 4-level ordinal cognition at 12 months
8. Stacked distributions of mRS at 12 months
9. Graph of imaging outcomes at 12 months (based on Table 9) displaying new incident infarct or haemorrhage, change in WMH, microbleeds, atrophy by allocated group.

**Figure 1**. CONSORT diagram

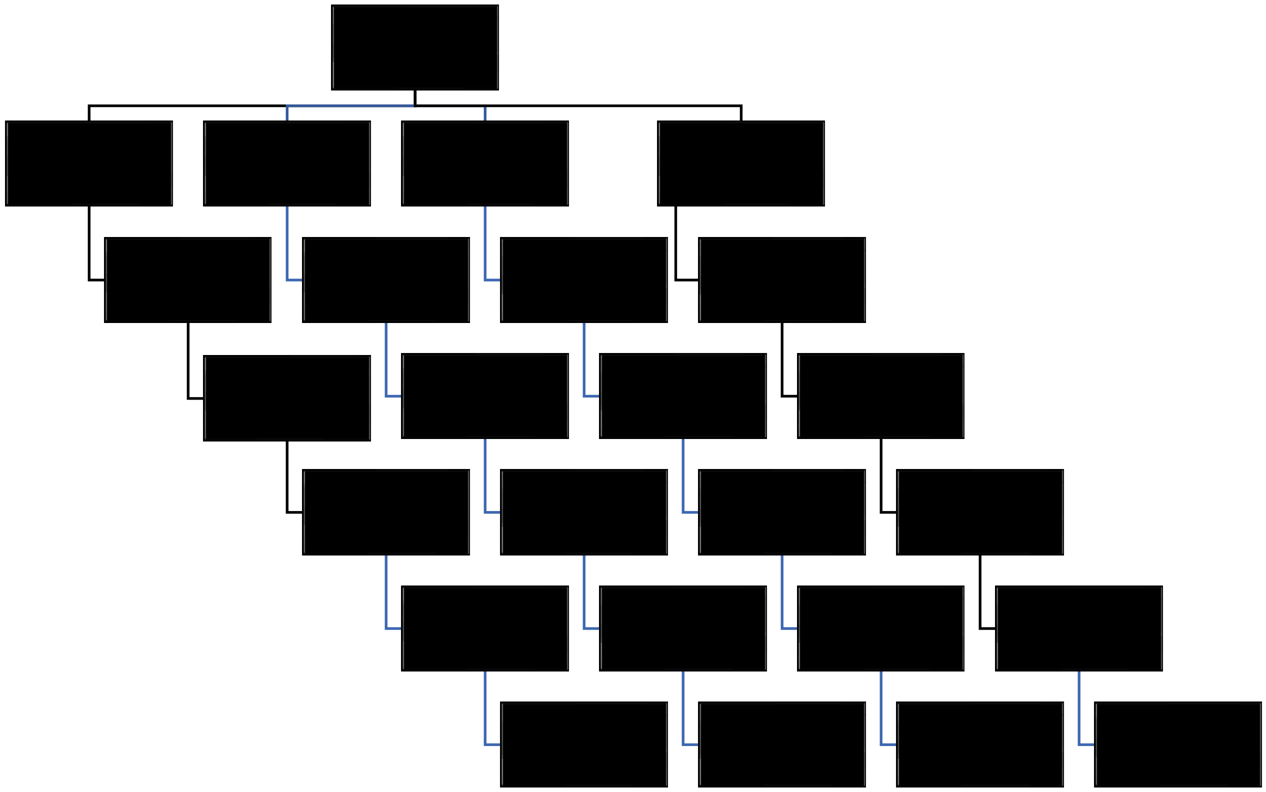

**Figure 2a**. Forest plot for isosorbide mononitrate versus none of composite outcome (stroke, TIA, MI, cognitive impairment, dependency or death) by clinical and adjudicated imaging subgroups. Adjusted

| | Interaction p |
|---|---|
| Age † | |
| <60 | - |
| 60-69 | - |
| ≥70 | - |
| Sex † | |
| Female | - |
| Male | - |
| Highest education † | |
| A-level (equivalent) or higher | - |
| O-level/GCSE (equivalent) | - |
| Primary or secondary primary school | - |
| Pre-morbid mRS † | |
| 0 | - |
| 1 | - |
| >1 | - |
| Time stroke-baseline † | |
| <180 days | |
| 180-364 days | |
| ≥365 days | |
| Smoking | |
| Never | |
| Past | |
| Present | |
| Hypertension | |
| No | - |
| Yes | - |
| Diabetes mellitus | |
| No | - |
| Yes | - |
| History of stroke or TIA | |
| No | - |
| Yes | - |
| Systolic BP † | |
| <130 | - |
| 130-159 | - |
| ≥160 | - |
| NIHSS † | |
| 0/1 | - |
| >1 | - |
| Relevant acute infarct on imaging | |
| No | - |
| Yes | - |
| Lacune on imaging | |
| Not visible | - |
| Visible | - |
| WMH or hypoattenuation | |
| Not visible | - |
| Visible | - |

| | |
|---|---|
| SVD score | |
| 0 | - |
| 1 | - |
| >1 | - |

† Minimisation variable

**Figure 2b**. Forest plot for cilostazol versus none of composite outcome (stroke, TIA, MI, cognitive impairment, dependency or death) by clinical and adjudicated imaging subgroups. Adjusted

| | Interaction p |
|---|---|
| Age † | |
| <60 | - |
| 60-69 | - |
| ≥70 | - |
| Sex † | |
| Female | - |
| Male | - |
| Highest education † | |
| A-level (equivalent) or higher | - |
| O-level/GCSE (equivalent) | - |
| Primary or secondary primary school | - |
| Pre-morbid mRS † | |
| 0 | - |
| 1 | - |
| >1 | - |
| Time stroke-baseline † | |
| <180 days | |
| 180-364 days | |
| ≥365 days | |
| Smoking | |
| Never | |
| Past | |
| Present | |
| Hypertension | |
| No | - |
| Yes | - |
| Diabetes mellitus | |
| No | - |
| Yes | - |
| History of stroke or TIA | |
| No | - |
| Yes | - |
| Systolic BP † | |
| <130 | - |
| 130-159 | - |
| ≥160 | - |
| NIHSS † | |
| 0/1 | - |
| >1 | - |
| Relevant acute infarct on imaging | |
| No | - |
| Yes | - |
| Lacune on imaging | |
| Not visible | - |
| Visible | - |
| WMH or hypoattenuation | |
| Not visible | - |
| Visible | - |
| SVD score | |

| | |
|---|---|
| 0 | - |
| 1 | - |
| >1 | - |

† Minimisation variable

**Figure 2c**. Forest plot for combined isosorbide mononitrate and cilostazol versus neither of composite outcome (stroke, TIA, MI, cognitive impairment, dependency or death) by clinical and adjudicated imaging subgroups. Adjusted

| | Interaction p |
|---|---|
| Age † | |
| <60 | - |
| 60-69 | - |
| ≥70 | - |
| Sex † | |
| Female | - |
| Male | - |
| Highest education † | |
| A-level (equivalent) or higher | - |
| O-level/GCSE (equivalent) | - |
| Primary or secondary primary school | - |
| Pre-morbid mRS † | |
| 0 | - |
| 1 | - |
| >1 | - |
| Time stroke-baseline † | |
| <180 days | |
| 180-364 days | |
| ≥365 days | |
| Smoking | |
| Never | |
| Past | |
| Present | |
| Hypertension | |
| No | - |
| Yes | - |
| Diabetes mellitus | |
| No | - |
| Yes | - |
| History of stroke or TIA | |
| No | - |
| Yes | - |
| Systolic BP † | |
| <130 | - |
| 130-159 | - |
| ≥160 | - |
| NIHSS † | |
| 0/1 | - |
| >1 | - |
| Relevant acute infarct on imaging | |
| No | - |
| Yes | - |
| Lacune on imaging | |
| Not visible | - |
| Visible | - |
| WMH or hypoattenuation | |
| Not visible | - |
| Visible | - |
| SVD score | |

| | |
|---|---|
| 0 | - |
| 1 | - |
| >1 | - |

† Minimisation variable

**Figure 3a**. Forest plot of 7 level cognition scale for isosorbide mononitrate versus none at 12 months by clinical and adjudicated imaging subgroups. Adjusted

| | Interaction p |
|---|---|
| Age † | |
| <60 | - |
| 60-69 | - |
| ≥70 | - |
| Sex † | |
| Female | - |
| Male | - |
| Highest education † | |
| A-level (equivalent) or higher | - |
| O-level/GCSE (equivalent) | - |
| Primary or secondary primary school | - |
| Pre-morbid mRS † | |
| 0 | - |
| 1 | - |
| >1 | - |
| Time stroke-baseline † | |
| <180 days | |
| 180-364 days | |
| ≥365 days | |
| Smoking | |
| Never | |
| Past | |
| Present | |
| Hypertension | |
| No | - |
| Yes | - |
| Diabetes mellitus | |
| No | - |
| Yes | - |
| History of stroke or TIA | |
| No | - |
| Yes | - |
| Systolic BP † | |
| <130 | - |
| 130-159 | - |
| ≥160 | - |
| NIHSS † | |
| 0/1 | - |
| >1 | - |
| Relevant acute infarct on imaging | |
| No | - |
| Yes | - |
| Lacune on imaging | |
| Not visible | - |
| Visible | - |
| WMH or hypoattenuation | |
| Not visible | - |
| Visible | - |
| SVD score | |

| | |
|---|---|
| 0 | - |
| 1 | - |
| >1 | - |

† Adjustment variable

**Figure 3b**. Forest plot of 7 level cognition scale for cilostazol versus none at 12 months by clinical and adjudicated imaging subgroups. Adjusted

| | Interaction p |
|---|---|
| Age † | |
| <60 | - |
| 60-69 | - |
| ≥70 | - |
| Sex † | |
| Female | - |
| Male | - |
| Highest education † | |
| A-level (equivalent) or higher | - |
| O-level/GCSE (equivalent) | - |
| Primary or secondary primary school | - |
| Pre-morbid mRS † | |
| 0 | - |
| 1 | - |
| >1 | - |
| Time stroke-baseline † | |
| <180 days | |
| 180-364 days | |
| ≥365 days | |
| Smoking | |
| Never | |
| Past | |
| Present | |
| Hypertension | |
| No | - |
| Yes | - |
| Diabetes mellitus | |
| No | - |
| Yes | - |
| History of stroke or TIA | |
| No | - |
| Yes | - |
| Systolic BP † | |
| <130 | - |
| 130-159 | - |
| ≥160 | - |
| NIHSS † | |
| 0/1 | - |
| >1 | - |
| Relevant acute infarct on imaging | |
| No | - |
| Yes | - |
| Lacune on imaging | |
| Not visible | - |
| Visible | - |
| WMH or hypoattenuation | |
| Not visible | - |
| Visible | - |
| SVD score | |

| | |
|---|---|
| 0 | - |
| 1 | - |
| >1 | - |

† Adjustment variable

**Figure 3c**. Forest plot of 7 level cognition scale for combined isosorbide mononitrate and cilostazol versus neither at 12 months by baseline subgroups. Adjusted

| | Interaction p |
|---|---|
| Age † | |
| <60 | - |
| 60-69 | - |
| ≥70 | - |
| Sex † | |
| Female | - |
| Male | - |
| Highest education † | |
| A-level (equivalent) or higher | - |
| O-level/GCSE (equivalent) | - |
| Primary or secondary primary school | - |
| Pre-morbid mRS † | |
| 0 | - |
| 1 | - |
| >1 | - |
| Time stroke-baseline † | |
| <180 days | |
| 180-364 days | |
| ≥365 days | |
| Smoking | |
| Never | |
| Past | |
| Present | |
| Hypertension | |
| No | - |
| Yes | - |
| Diabetes mellitus | |
| No | - |
| Yes | - |
| History of stroke or TIA | |
| No | - |
| Yes | - |
| Systolic BP † | |
| <130 | - |
| 130-159 | - |
| ≥160 | - |
| NIHSS † | |
| 0/1 | - |
| >1 | - |
| Relevant acute infarct on imaging | |
| No | - |
| Yes | - |
| Lacune on imaging | |
| Not visible | - |
| Visible | - |
| WMH or hypoattenuation | |
| Not visible | - |
| Visible | - |
| SVD score | |

| | |
|---|---|
| 0 | - |
| 1 | - |
| >1 | - |

† Adjustment variable

**Figure 4a**. Forest plot of Wei-Lachin global outcome for isosorbide mononitrate versus none by subgroups. Adjusted

| | Interaction p |
|---|---|
| Age † | |
| <60 | - |
| 60-69 | - |
| ≥70 | - |
| Sex † | |
| Female | - |
| Male | - |
| Highest education † | |
| A-level (equivalent) or higher | - |
| O-level/GCSE (equivalent) | - |
| Primary or secondary primary school | - |
| Pre-morbid mRS † | |
| 0 | - |
| 1 | - |
| >1 | - |
| Time stroke-baseline † | |
| <180 days | |
| 180-364 days | |
| ≥365 days | |
| Smoking | |
| Never | |
| Past | |
| Present | |
| Hypertension | |
| No | - |
| Yes | - |
| Diabetes mellitus | |
| No | - |
| Yes | - |
| History of stroke or TIA | |
| No | - |
| Yes | - |
| Systolic BP † | |
| <130 | - |
| 130-159 | - |
| ≥160 | - |
| NIHSS † | |
| 0/1 | - |
| >1 | - |
| Relevant acute infarct on imaging | |
| No | - |
| Yes | - |
| Lacune on imaging | |
| Not visible | - |
| Visible | - |
| WMH or hypoattenuation | |
| Not visible | - |
| Visible | - |
| SVD score | |

| | |
|---|---|
| 0 | - |
| 1 | - |
| >1 | - |

† Adjustment variable

**Figure 4b**. Forest plot of Wei-Lachin global outcome for cilostazol versus none by subgroups. Adjusted

| | Interaction p |
|---|---|
| Age † | |
| <60 | - |
| 60-69 | - |
| ≥70 | - |
| Sex † | |
| Female | - |
| Male | - |
| Highest education † | |
| A-level (equivalent) or higher | - |
| O-level/GCSE (equivalent) | - |
| Primary or secondary primary school | - |
| Pre-morbid mRS † | |
| 0 | - |
| 1 | - |
| >1 | - |
| Time stroke-baseline † | |
| <180 days | |
| 180-364 days | |
| ≥365 days | |
| Smoking | |
| Never | |
| Past | |
| Present | |
| Hypertension | |
| No | - |
| Yes | - |
| Diabetes mellitus | |
| No | - |
| Yes | - |
| History of stroke or TIA | |
| No | - |
| Yes | - |
| Systolic BP † | |
| <130 | - |
| 130-159 | - |
| ≥160 | - |
| NIHSS † | |
| 0/1 | - |
| >1 | - |
| Relevant acute infarct on imaging | |
| No | - |
| Yes | - |
| Lacune on imaging | |
| Not visible | - |
| Visible | - |
| WMH or hypoattenuation | |
| Not visible | - |
| Visible | - |
| SVD score | |

| | |
|---|---|
| 0 | - |
| 1 | - |
| >1 | - |

† Adjustment variable

**Figure 4c**. Forest plot of Wei-Lachin global outcome for combined isosorbide mononitrate and cilostazol versus neither by subgroups. Adjusted

| | Interaction p |
|---|---|
| Age † | |
| <60 | - |
| 60-69 | - |
| ≥70 | - |
| Sex † | |
| Female | - |
| Male | - |
| Highest education † | |
| A-level (equivalent) or higher | - |
| O-level/GCSE (equivalent) | - |
| Primary or secondary primary school | - |
| Pre-morbid mRS † | |
| 0 | - |
| 1 | - |
| >1 | - |
| Time stroke-baseline † | |
| <180 days | |
| 180-364 days | |
| ≥365 days | |
| Smoking | |
| Never | |
| Past | |
| Present | |
| Hypertension | |
| No | - |
| Yes | - |
| Diabetes mellitus | |
| No | - |
| Yes | - |
| History of stroke or TIA | |
| No | - |
| Yes | - |
| Systolic BP † | |
| <130 | - |
| 130-159 | - |
| ≥160 | - |
| NIHSS † | |
| 0/1 | - |
| >1 | - |
| Relevant acute infarct on imaging | |
| No | - |
| Yes | - |
| Lacune on imaging | |
| Not visible | - |
| Visible | - |
| WMH or hypoattenuation | |
| Not visible | - |
| Visible | - |
| SVD score | |

| | |
|---|---|
| 0 | - |
| 1 | - |
| >1 | - |

† Adjustment variable

**Figure 5**. Forest plot of central imaging reads as per list for table 9 of absolute and change in imaging findings

a) Isosorbide mononitrate versus none
b) Cilostazol versus none
c) Combined isosorbide mononitrate and cilostazol versus neither

**Figure 6**. Stacked distributions of 7-level ordinal cognition at 12 months

d) Isosorbide mononitrate versus none
e) Cilostazol versus none
f) Combined isosorbide mononitrate and cilostazol versus neither

**Figure 7**. Stacked distributions of 4-level ordinal cognition at 12 months

a) Isosorbide mononitrate versus none
b) Cilostazol versus none
c) Combined isosorbide mononitrate and cilostazol versus neither

**Figure 8**. Stacked distributions of mRS at 12 months

a) Isosorbide mononitrate versus none
b) Cilostazol versus none
c) Combined isosorbide mononitrate and cilostazol versus neither

**Figure 9**. Graph of imaging outcomes at 12 months (based on Table 9) displaying new incident infarct or haemorrhage, change in WMH, microbleeds, atrophy by allocated group.

a) Isosorbide mononitrate versus none
b) Cilostazol versus none
c) Combined isosorbide mononitrate and cilostazol versus neither

**Signature:**

### LACI-2 SAP v1.5 20220606

Final Audit Report 2022-06-15

| | |
|---|---|
| Created: | 2022-06-08 |
| By: | Sian Irvine |
| Status: | Signed |
| Transaction ID: | CBJCHBCAABAAhjIv2n-mGfbkbnDio0Cvi-NrpFWzqFO9 |

#### "LACI-2 SAP v1.5 20220606" History

Document created by Sian Irvine

2022-06-08 - 12:51:32 PM GMT- IP address: 31.48.48.183

2022-06-08 - 12:53:48 PM GMT

2022-06-09 - 9:01:11 AM GMT- IP address: 92.40.132.99

Document e-signed by Philip Bath

Signature Date: 2022-06-09 - 9:02:15 AM GMT - Time Source: server- IP address: 92.40.132.99

2022-06-09 - 9:02:19 AM GMT

2022-06-09 - 11:04:31 AM GMT- IP address: 172.224.227.28

Document e-signed by Joanna Wardlaw

Signature Date: 2022-06-09 - 1:31:46 PM GMT - Time Source: server- IP address: 212.187.183.33

2022-06-09 - 1:31:49 PM GMT

2022-06-15 - 7:54:36 AM GMT- IP address: 167.98.250.154

Document e-signed by alan montgomery

Signature Date: 2022-06-15 - 7:55:08 AM GMT - Time Source: server- IP address: 167.98.250.154

2022-06-15 - 7:55:11 AM GMT

2022-06-15 - 7:58:40 AM GMT- IP address: 129.215.89.68

A signed copy was uploaded by Sian Irvine

2022-06-15 - 9:26:35 AM GMT

Agreement completed.

2022-06-15 - 9:26:35 AM GMT
